# Navigating the DiGA Jungle: A Taxonomy and Archetypal Framework of the German Digital Therapeutics Landscape

**DOI:** 10.64898/2025.12.30.25343225

**Authors:** Egidia Cenko, Thure Georg Weimann, Georgios Raptis

**Author notes:** These authors contributed equally to this work (Egidia Cenko, Thure Georg Weimann).

## Abstract

Digital therapeutics (DTx) are patient-facing apps designed to support individuals in their daily lives. Therefore, they have the potential to revolutionize healthcare by empowering and engaging patients to become active players in their own care. Despite the increasing adoption of DTx in national healthcare systems, research on their design remains limited. The present study introduces “DiGATax”, a taxonomy designed to categorize and analyze DTx, including perspectives on content, intervention delivery logic and technology, as well as the patient’s interface, consolidating and expanding upon prior taxonomic work. Based on *n* = 44 applications retrieved from the German DiGA directory that demonstrated positive health outcomes, the taxonomy is supported by empirical evidence. Additionally, the study contributes by presenting an archetype framework of DTx derived from a taxonomy-based cluster analysis. Further analyses offer insights into specific combinations of DTx characteristics across archetypes, the user interface as a key factor in their acceptance, and potential links between DTx design and health-related and user engagement outcomes. By offering new insights into DTx design, this study contributes towards more organized research and reporting, ultimately paving the way for the development of effective solutions. It also marks a further step towards Meta-DTx, which aim to align patient care for multimorbid patients under one umbrella.

## 1 Introduction

The digital transformation in healthcare comes with innovations and changes for various players [1]. An important trend that emerges from this is the shift in the role of patients towards becoming active members of the care team, helping to shape their own patient journey and being recognized as key decision-makers [2]. It is well documented that patient-centered care can enhance empowerment and engagement of patients, leading to improved health outcomes and cost savings [3, 4]. For instance, it has been shown that self-management programs targeting behavioral modifications for patients with chronic diseases (e.g., type 2 diabetes, obesity, or mental illness) have been effective to promote physical activity, health responsibility, and overall outcomes [5].

As a result of the digital transformation, self-management interventions are gradually being supported with patient-facing solutions that are integrated into everyday life, successively reshaping care pathways [6]. Notably, the trend toward patient (or “consumer”)-directed information systems that support the individual’s self-care of a disease was even forecasted more than two decades ago by Eysenbach [7], with the introduction of DTx marking a big leap forward. In general, a medical indication, usually defined by the International Classification of Diseases, serves as the foundation for using DTx [8]. According to the International Organization for Standardization (ISO), DTx can be defined as “health software intended to treat or alleviate a disease, disorder, condition, or injury by generating and delivering a medical intervention that has a demonstrable positive therapeutic impact on a patient’s health” [9]. While this definition is quite broad, they are establishing themselves in particular as a new therapeutic option or complement to the treatment of behavior-modifiable diseases [10, 11, 12].

Despite the great potential of digital behavior change interventions, including DTx, long-term evidence is up to now still scarce [13, 14]. One reason discussed in the literature is that interventions often focus on deliberative psychological processes to adopt new behaviors, while digitally supported guidance for transforming them into less-or non-conscious automated behaviors (i.e., habits) is limited [15, 16]. Although techniques related to habit formation are recognized in conceptual frameworks, the terminology differs between authors [15, 17]. In addition, behavioral changes serve a dual purpose in DTx. On the one hand, they can be part of the “digital pill” to change behavior in such a way that it has a positive effect on the disease. On the other hand, “Apps don’t work for patients who don’t use them” as recently stated by Schwartz et al. [18], underscoring the need to promote engagement behavior in relation to DTx themselves.

One factor contributing to the increasing implementation of DTx are regulatory frameworks that allow them to be prescribed and reimbursed like traditional pharmaceuticals [10]. While Germany lagged behind other European countries with the introduction of a national electronic health record [19], it has been a forerunner in the field of DTx for several years and a role model for other countries [20]. Their DTx implementation, namely “DiGAs” (Digitale Gesundheitsanwendungen), follows the mandatory requirements set by BfArM, the German regulatory authority for medicinal products and medical devices. In addition to Germany, the United Kingdom, Belgium, France and Spain were also early adopters in Europe [21], with Korea and China standing out internationally [22].

Through the rise of DTx, new opportunities for research emerged to enhance the understanding of aspects related to data, their integration into social processes, their technological design, and their value impact as outlined by Fürstenau et al. [23]. For guiding the future development of effective DTx, systematically describing, analyzing and understanding the current state-of-the art is essential. Previous studies examined non-technical factors in the approval process, such as the clinical study design and evidence [8, 24], alongside economic analyses with regard to pricing and cost-effectiveness [25], as well as adoption factors including prescriptions and technological acceptance [26, 20] for DiGAs in Germany. However, studies on systematically describing and analyzing the content-related and technical aspects of DTx are still scarce. This need is further underscored when considering the researchers’ claim that standardized frameworks are needed to guide the development and evaluation of DTx [27, 28]. The need for greater standardization is further emphasized, as current evaluation outcomes focus predominantly on health-related improvements without being able to attribute them to integrated features due to the lack of systematization of existing DTx [29, 30]. Similar issues have been also described in the broader field of telemedicine with the demand for more taxonomical guidance [31]. In general, taxonomies provide a systematic classification of certain objects of interest and have been established as a basis for conceptualizing undiscovered phenomena, allowing a structured approach to understand new innovations [32].

During the preparation of this manuscript, the study by Weimar et al. [33] was published, providing first insights into features of DiGAs, especially those visible at the interface level. While their work is a first step to address this gap and supports our argument, it is limited by its purely inductive approach. In particular, it does not build on existing taxonomic research in adjacent fields and also does not intend to present a taxonomy in the sense of Nickerson et al. [34] with clearly described dimensions and characteristics. Their work focuses on a frequency-based analysis of features at the overall level without aiming to explore subtypes (clusters) of DTx. Furthermore, by focusing on the visual features, it does not investigate aspects related to the intervention delivery logic, as well as the actual “active ingredients” which typically manifest as behavior change techniques (BCTs) at the content level [35]. As a result, only a subset of the DTx design space has been investigated yet, providing starting points to build on and extend their work. Moreover, their framework includes aspects like data security measures, which are certainly relevant but may be less suitable as a differentiating dimension in a taxonomy. For example, data security requirements are predominantly shaped by regulatory frameworks, which are subject to continuous revision and apply uniformly across all DiGAs. Consequently, such dimensions may be regarded as general prerequisites rather than criteria that meaningfully distinguish between different DTx applications.

The present study seeks to offer new perspectives in different ways. First, it aims to systematically develop a comprehensive taxonomy including dimensions and characteristics related to i) the content, ii) intervention delivery logic, as well as iii) the patient’s interface. Consequently, a holistic view on DTx should be taken into account, allowing a systematic classification of design-related differences. For this purpose, both a conceptual-to-empirical as well as an empirical-to-conceptual approach, as outlined by the method by Nickerson et al. [34], are followed to ensure alignment with previous taxonomical work and derive new extensions. In accordance with Weimar et al. [33], we also focus on permanently listed DiGAs from the German directory [36], as they have passed the rigorous approval process and demonstrated significant positive effects on patient care.

Second, using the developed taxonomy and set of categorized DTx applications, groups of design patterns should be explored through hierarchical clustering, referred to as “archetypes”. Based on the clustering results, an archetype framework is developed and evaluated using test objects (DTx applications) that were not involved in the initial taxonomy development and clustering process.

Third, the present work aims to investigate detailed combinations regarding BCTs as the active ingredients, the composition and navigational aspects of the user interface (UI) as well as associations to intervention outcomes. The latter is based on the idea that different targeted outcomes might call for different DTx designs, although this is not yet fully understood [37]. By basing our work on clinically evaluated interventions that have already proven effective, we aim to ensure the “higher-quality evidence” as demanded by Mair et al. [37]. Furthermore, engagement was found to be a decisive factor regarding the overall effectiveness and is hypothesized to moderate the effects between intervention design and outcome according to Perski et al. [35]. Using publicly available user ratings in the app store, the present study also aims to exploratively investigate discriminative associations between ratings and DTx characteristics that could serve as a starting point for new hypotheses.

To address the outlined goals, the following research questions (RQs) should be explored, as shown in the conceptual research model of this study in Figure 1:

**Figure 1:**
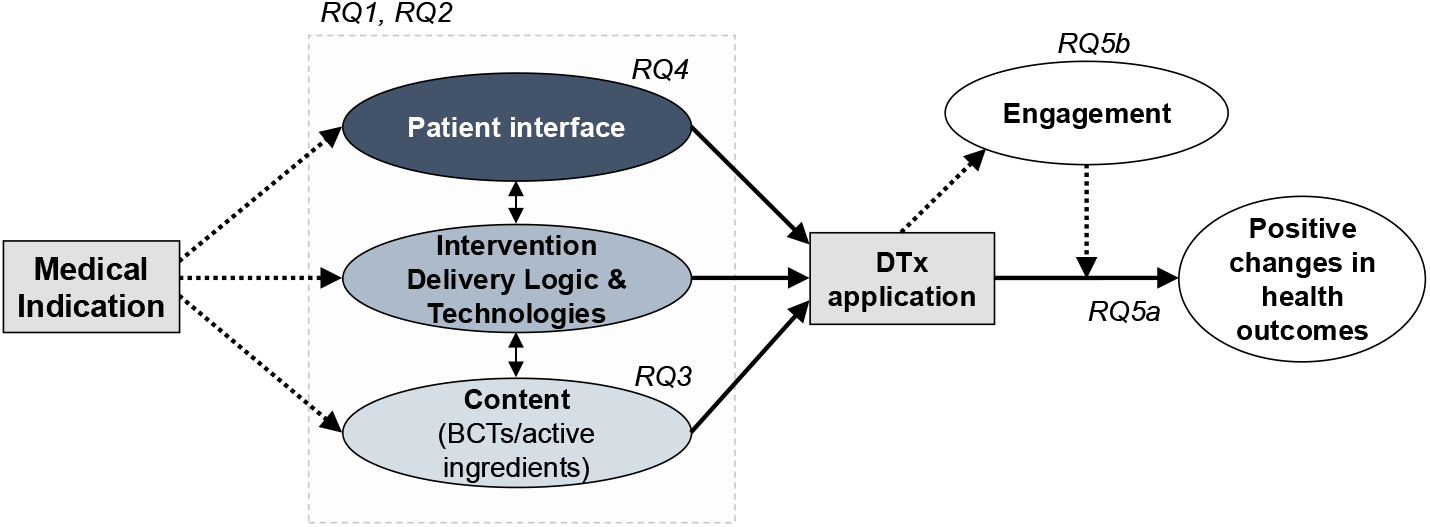
Conceptual research model with interrelationships and marked research questions.nn

**RQ1:** *What are dimensions and characteristics of DTx interventions related to the content, intervention delivery logic and technology, as well as the patient’s interface?*

**RQ2:** *Which archetypes can be identified in the current German DTx landscape?*

**RQ3:** *What combinations of “active ingredients” (BCTs) are frequently used?*

**RQ4:** *How is the user interface of DTx interventions typically structured regarding navigational aspects and functional areas?*

**RQ5:** *Which potential associations can be identified between the DTx design and* ***(a)*** *health-related outcomes as well as* ***(b)*** *user engagement?*

The remainder of this paper is organized as follows: the next section details our method, then section three presents our results in dedicated subsections addressing the stated research questions. The article concludes with a discussion of future directions for research and practice in the field of DTx.

## 2 Method

To comprehensively address the stated research goals and questions, we followed a multi-stage process. Initially, we conducted a systematic literature review (step 1) to identify potential dimensions and characteristics underlying DTx applications. Therefore, we searched for existing taxonomies in related fields, such as telemedicine, digital behavior change and the field of mobile health (“mHealth”), which could provide a conceptual foundation for developing a dedicated taxonomy for DTx. The systematic review of the literature followed the PRISMA selection flow [38], and the results were organized according to the concept-centric approach described by Webster and Watson [39] (step 2). Building on the systematically structured results of the literature review, we started with the taxonomy development process following the method established by Nickerson et al. [34]. As recommended in the subsequent work “An update for taxonomy designers” by Kundisch et al. [40], we conducted both a conceptual-to-empirical and empirical-to-conceptual approach (step 3). Likewise, we used new objects that became listed as permanent after analyzing the first set of *n* = 35 objects as of July 2024, to evaluate the comprehensiveness and applicability of the preliminary taxonomy and identify potential refinements. In total, *n* = 9 additional DiGAs were retrieved one year later, by the end of July 2025, and used for evaluation. To uncover DTx archetypes, we employed a taxonomy-based clustering analysis (step 4) on the *n* = 35 applications and also evaluated the archetypes based on the additional objects. We have considered the recommendations for taxonomy-based data clustering by Heumann et al. [41] for general methodological guidance in this regard. Steps 5 and 6 involved in-depth analyses of specific combinations of characteristics across the identified archetypes and at the overall level. To provide further exploratory insights, the outcome perspective was added in terms of outcome categories and user ratings as an operationalization of engagement. Figure 2 depicts the overall methodological process, with the following subsections providing further details on our pursued approach.

**Figure 2:**
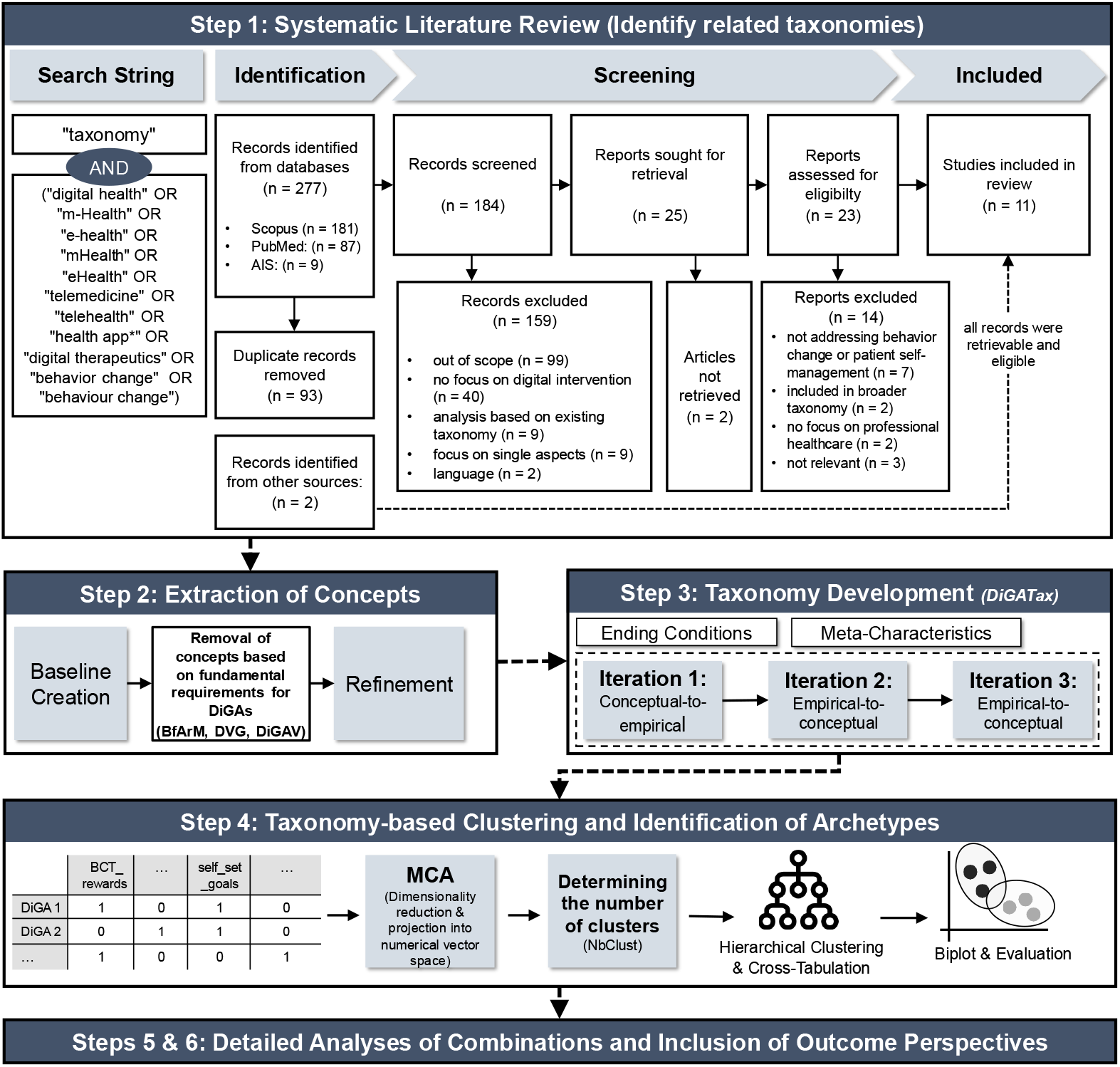
Overview of the methodological approach.

### 2.1 Systematic literature review (step 1)

For the literature review, we considered the databases PubMed and Scopus to include articles related to both medicine and technology, providing a representative basis for the target domain. Considering the origin of the seminal work on taxonomy development by Nickerson et al. [34] in information systems research, we decided to extend our search also to the corresponding research community using the AIS eLibrary. Furthermore, our literature search was complemented by a semi-systematic search using the search terms below in Google and Google Scholar.

Our search string concatenated the term “taxonomy” (title) with relevant strings in the field of behavior change and digital health interventions (title and abstract; in Scopus also keywords) (see Figure 2). The initial literature search was conducted in May 2024 and repeated on September 10, 2025. Upon repeating the search, we identified two additional papers that met the criteria stated below. However, as the dimensions and/or characteristics with relevance to the meta-characteristics of our work were already reflected in our taxonomy at this stage, no changes occurred. Nonetheless, we aligned the corresponding dimensions and/or characteristics and referenced the papers for completeness (see appendix A).

The search yielded 277 results and *n* = 184 papers remained after duplicate removal (September 2025). The inclusion and exclusion criteria for the screening step were agreed upon jointly by the authors. Our inclusion criteria were defined to consider papers that i) specifically focus on creating a taxonomy for digital health interventions and related sub fields *and* ii) integrating technical aspects as well as aspects related to the patient’s self-management (including behavior change). Our major focus was on taxonomies that provide a holistic view on patient-facing interventions in professional care settings. However, we decided to extend our consideration to taxonomies that concentrate on specific “micro”-aspects related to the design, such as only gamification, when the broader taxonomies lack sufficient detail and the analysis material clearly indicates that further aspects are essential.

Overall, we excluded papers that: i) did not focus on digital health or digital behavior change with a therapeutic goal, ii) only combined or used other identified taxonomies without empirical extensions, and/or iii) were written in a language other than English or German. Additionally, iv) taxonomies that were used to derive a more comprehensive taxonomy in a subsequent work were also excluded, and only the most comprehensive one was used. Papers were also excluded when institutional access was not possible (v). Notably, of those papers that analyzed digital interventions based on an existing taxonomy, 8 of 9 papers used the behavior change technique taxonomy (BCTT) by Michie et al. [17], highlighting its widespread use for analyzing the active ingredients.

The initial literature screening and analysis were conducted by one author, while a second author repeated the literature search, reviewed and validated the coded dimensions as well as characteristics across the identified works. In sum, our literature search in databases resulted in *n* = 9 taxonomies and was extended by *n* = 2 taxonomies/frameworks identified via the other search engines.

### 2.2 Extraction of concepts (step 2)

The results of the literature review were used to analyze existing taxonomies with respect to their dimensions and characteristics. This extraction and synthesis step was particularly guided by our perspectives of interest (meta-characteristics, see step 3). In addition, as our work focuses on the German DTx realization, we analyzed the preliminary concepts of the technical aspects and compared them to the requirements defined by German law (DiGAV and DVG) and the BfArM guidelines [42]. This was done to directly exclude fundamental concepts that are expected by law in all listed DiGAs (e.g., data security, interoperability) and that will therefore not vary among the analyzed applications. For reference, a mapping of how the final taxonomy aligns with the dimensions and characteristics proposed in prior studies is included in the appendix A.

### 2.3 Taxonomy development (step 3)

Following the method by Nickerson et al. [34], the definition of the meta-characteristic represents the starting point. For the present work we considered the meta-characteristic as the systematic classification of DTx concerning content, intervention delivery logic and technologies, as well as the patient’s interface. Subsequently, the method requires the definition of ending conditions. With regard to the objective ending conditions, we generally followed the eight mentioned by Nickerson et al. [34]. However, the strict definition of mutual exclusivity is extended to also allow non-mutually exclusive characteristics within a dimension. In our work, forcing mutual exclusiveness per dimension might not reflect the complexity of DTx, thus threatening the purpose of the taxonomy. Möller et al. [43], recommend a clear indication of exclusivity for the field of business model design, i.e., dimensions that adopt complete or non-mutual exclusiveness are required to be denoted as such. Likewise, Strobel et al. deliberately included non-mutually exclusive characteristics to enhance readability [44]. Overall, their proposal is adopted for our case to better reflect the complexity, improve readability, and thus also enhance the taxonomy’s applicability. Last but not least, the subjective ending conditions comprehensiveness and robustness are addressed by analyzing an additional set of “new” DiGAs that were not used for the development of the taxonomy before (investigating completeness), as well as the interpretation of the cluster analysis (investigating distinctiveness).

Using the results from step 2, we first employed the conceptual-to-empirical path (iteration 1). Subsequently, the iterations 2 and 3 followed the empirical-to-conceptual approach where the DTx (DiGA) applications were examined in detail. In accordance with the work by Weimar et al. [33], our analysis was primarily based on the available information sources from the DiGA directory and included i) manufacturer descriptions, ii) information for healthcare professionals (HCPs), iii) instructions and manuals provided by the manufacturer, iv) their linked website v) the linked application (depending on the available application type, information in the Google Play Store, Apple App Store and/or on the actual DTx website). However, we also considered information retrieved in scientific and gray publications by the DiGA provider. Furthermore, when screenshot material was insufficient for coding, test versions by the DiGA providers or insights from “DigaDocs” [45] were used (if available) to obtain an accurate categorization. DigaDocs aims to provide independent and reliable information for both patients and HCPs by “hands on” testing DiGAs.

Based on a preliminary draft of the taxonomy elaborated by one author, two authors analyzed the applications, mapped them to the identified characteristics or conducted refinements when necessary. Inter-rater reliability was evaluated on the first set of *n* = 35 applications using Cohen’s *κ* statistic, yielding a value of 0.836, which indicated strong agreement between the coders [46]. Disagreements were discussed and resolved collaboratively as expert consensus. The *n* = 9 additional applications were coded by one author and minor refinements or extensions to the definitions of the dimensions and characteristics were discussed together. Finally, the taxonomy was collaboratively reviewed for redundancies, and related characteristics or dimensions were merged with others and/or removed.

### 2.4 Taxonomy-based clustering and evaluation of archetypes (step 4)

Using the binary coded data, we applied a cluster analysis to identify existing DiGA groups that share similar characteristics. All subsequent reported analyses were performed using the statistical software R (v4.5.1). Considering the high dimensionality of the taxonomy with 108 characteristics (representing binary variables), dimensionality reduction is critical before applying distance-based similarity measures as part of the clustering process. As outlined by Heumann et al. [41], a frequently observed methodical issue in taxonomy research is the application of common clustering algorithms to categorical data. To address both this issue and the high dimensionality of our data, we first conducted a multiple correspondence analysis (MCA) using the R package called `FactoMineR` [47]. We retained the first 16 dimensions that cumulatively accounted for 85% of the explained variance.

As we also aim to understand nested structures among the clusters, we decided to employ hierarchical clustering using the established Ward’s method as implemented in R. Following the recommendations by Heumann et al. [41], we utilized quantitative metrics to determine a reasonable number of clusters. In particular, we used the R package `NbClust` that computes multiple metrics in parallel, including the silhouette score, gap statistic or Davies-Bouldin score, and uses a majority vote for a suggested number of clusters [48]. Given the relatively small data set, we defined three clusters as the minimum and seven clusters as the maximum size to still enable a meaningful interpretation (i.e., on average at least 5 DiGAs per cluster). Furthermore, we defined as a criterion that no archetype cluster should include more than half of the number of objects. After the determination of the number of clusters, a cross tabulation analysis was conducted based on the distributions of the observed dimensions over the clusters.

To understand the archetypes and their relationships, we investigated the dendrogram and visualized the first two dimensions of the MCA model. Subsequently, we evaluated the robustness and comprehensiveness of the archetypes using the *n* = 9 additional DiGAs by projecting them into the two-dimensional space. Therefore, we first fitted an MCA model using the entire data set (*n* = 44, “training” and “test” set) and retained the first 16 dimensions. Subsequently, we employed the *k*-nearest neighbors algorithm (kNN) using the euclidean distance to identify the six closest DTx applications in the neighborhood. We chose *k* = 6 by applying the heuristic of selecting the number of nearest neighbors as the square root of the training set size 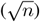 [49].

Then, we computed an interpolated projection of the additional DiGAs in this two-dimensional space using an average of the coordinates of the neighbors from the first MCA model (“training” set). This allows for the interpretation of the extent to which the new objects fit into the archetype framework. Following Szopinski et al. [50], this evaluation of the taxonomy and the resulting archetypes aligns with the approach of an “illustrative scenario” that is commonly used for this purpose.

### 2.5 Detailed analyses of combinations and inclusion of outcome perspectives (step 5 and 6)

Building on the coded objects along the taxonomy characteristics and the identified archetypes, we analyzed associations of characteristics using a graph-based network analysis [51] (step 5). In a first step, frequency matrices were calculated reflecting the co-occurrence of certain combinations of characteristics. Afterwards, these matrices were visualized as a network. We used the R package igraph [52] along with the “Louvain” community detection algorithm [53] to uncover more detailed groups of characteristics. Finally, we analyzed associations of the DTx design with respect to the targeted intervention outcome categories and user engagement (step 6). To categorize the intervention outcomes, the evaluation study designs of the included DiGAs were analyzed regarding their primary outcomes (reported in the DiGA directory). The categorization was then conducted based on the outcome domains described in the taxonomy by Dodd et al. [54] and the hierarchical taxonomy of psychopathology (“HiTOP”) [55] in a first step. To exploratory investigate the potential of the taxonomy to support hypothesis generation, we conducted a regression tree analysis based on the characteristics as predictors and the app store ratings as response variable. We used the R package `rpart` for regression tree modeling [56]. In line with prior research in the field of DiGAs [57], we used the app store ratings by users as a proxy measure for user engagement that can be defined as the user’s “subjective experience” according to Perski et al. [35]. We collected ratings from both the Google Play and Apple App Store, then calculated a weighted average based on the number of ratings in each store. However, as there were no comparable equivalents available for applications that are solely provided as web-apps, we needed to limit our analysis to those published via the app stores. In addition, two providers with multiple interventions for different medical indications published one “umbrella app” from which the corresponding intervention is then accessed. Since only one rating for multiple DiGA interventions is available in this case, we decided to use only the intervention with the most prescriptions to reduce bias. The number of prescriptions was retrieved from the most recent DiGA report published by the national association of statutory health insurances in Germany [58].

## 3 Results

### 3.1 Related taxonomies and extraction of concepts

The second step of our research process comprised the extraction, consolidation and systematization of dimensions and characteristics found in existing taxonomies. Overall, the taxonomies largely varied in their comprehensiveness and domain focus. They spanned a broad range, including taxonomies that aim to cover the entire field of digital health, published by the World Health Organization [59], as well as the subfields of telemedicine [31], mobile health (mHealth) [60, 61, 62], and specific application types such as patient portals [63]. With regard to mHealth also more specific taxonomies were retrieved focusing on particular medical indications, including COVID-19 [64], cancer care [65] and depression [66]. Furthermore, we identified a taxonomy focusing on self-management of diabetes covering both mHealth and telemedicine [67]. Although not designed and intended as a taxonomy in the sense of Nickerson et al. [34], we also considered the work by Weimar et al. [33] as a “taxonomy-like” framework that categorizes features. As mentioned in section 2.2, we excluded foundational characteristics (e.g., certification). In sum, we identified and consolidated *n* = 15 dimensions related to our meta-characteristics based on prior work. These were:

- **Patient interface:** *patient-generated health data* (*n* = 7), *communication* (*n* = 5), the modalities of *information provision* (*n* = 2), *menu type of UI* (*n* = 1)
- **Intervention delivery logic and technologies:** *technological infrastructure* (*n* = 4), *user* (*n* = 2), *community* (*n* = 2), *notification* (*n* = 2), *communication/information flow* (*n* = 2), *therapy type* (*n* = 2), *personalization approach* (*n* = 1), *intervention frequency* (*n* = 1), *intervention duration* (*n* = 1), *data connectivity* (*n* = 1)
- **Content:** *behavior change techniques* (*n* = 1)

### 3.2 Derived taxonomy (DiGATax)

Although several dimensions and characteristics of the identified taxonomies had an appropriate level of detail regarding our meta-characteristics (content, intervention delivery logic and technologies, as well as the patient’s interface), some aspects still required further refinement. Since the BCTT by Michie et al. [17] is widely recognized as a standard in the literature, we chose to align the content perspective closely with it. Given the importance of long-term maintenance of behavioral changes [68], we also aligned the BCTT with the habit-altering strategies outlined by Pinder et al. [15].

Employing the preliminary taxonomy based on the extracted concepts from prior work, we first conducted the conceptual-to-empirical step by coding all *n* = 35 applications along the derived dimensions and characteristics. As we observed during this step that existing DTx interventions are also incorporating gamified elements and more advanced algorithms (including AI), we decided to refine this conceptualization. Both concepts have not yet been represented as corresponding dimensions with multiple characteristics in the analyzed literature and were only found in 2 of the 11 articles as single characteristics (see [61, 67]). Regarding gamification, we expanded on the works by Schmidt-Kraepelin et al. [69] and Hervas et al. [70], which were initially excluded in our literature search due to their narrow focus. We focused on the dimensions goal setting (how it is realized) and user advancement from [69] that are both related to the intervention delivery logic perspective. The concrete realization of gamified design elements at the interface level was guided by the consolidated terminology by [70].

Regarding the use of advanced algorithms, we decided to derive characteristics during the empirical-to-conceptual steps due to the reliance on textual information from the DiGA provider, which can vary greatly in detail. Unlike gamified design elements, which are often recognizable through the user interface, intelligent algorithms are a more hidden feature, with related terms like “AI” may also be used as general trend terms. Notably, one dimension (“communication/information flow” [66, 62]) was dropped after this step, as we noticed that all existing applications can be regarded as interactive without focusing solely on information provision or data reporting. However, it served as inspiration, along with the dimension “user advancement”, by [69] for deriving the new dimension “intervention flow”. Overall, the second iteration resulted in four new dimensions (intervention flow, use of advanced algorithms, gameful design and functional areas). In short, “intervention flow” describes how the patient progresses through the therapeutic program (time-based, completion-based, time- and completion-based, path with unconstrained access). The dimension “use of advanced algorithms” refers to their purpose. This can be content personalization at the macro level (i.e., tailoring of the composition and chronological order of content) or micro level (i.e., tailoring within content blocks that are the same for multiple users, e.g., branching within an educational module). Further, this dimension comprises reactive contextual guidance (e.g., when emergency situations are detected), descriptive data analysis, and computer vision. The dimension gameful design subsumes the integration of game-like elements as part of the intervention, including badges, scores, levels and serious games. Last but not least, functional areas concentrate the software capabilities at the interface level (e.g., diary, program, conversation).

As part of the third iteration (empirical-to-conceptual), additional *n* = 9 DiGAs were analyzed. This resulted in only marginal adjustments or clarifications to the definitions of the characteristics and dimensions, with no new ones being added. Table 1 summarizes all 19 dimensions and 108 characteristics. Detailed descriptions can be found in appendix A.

**Table 1:**
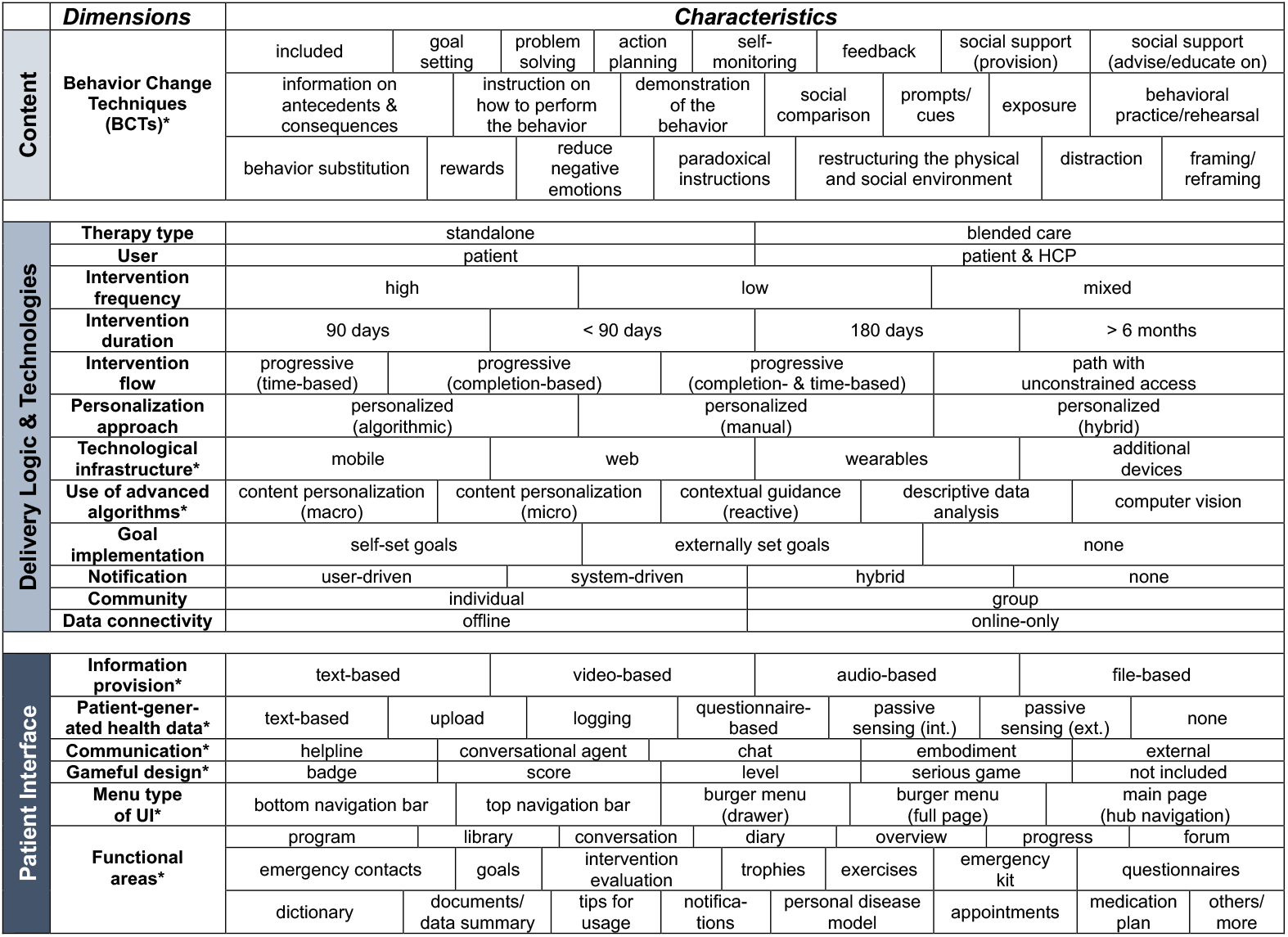
Final taxonomy “DiGATax” (* = non-mutually exclusive dimensions).

### 3.3 Cluster analysis and identification of archetypes

A cluster analysis was performed using the *n* = 35 coded applications after the second iteration. As described in section 2.4, we first conducted a dimensionality reduction using MCA, then employed NbClust to identify an appropriate number of clusters (Ward’s method (D2)). We limited the coding of the functional areas for clustering to those accessible at the first navigational level of the UI, as this usually reflects the primary functions and common usage patterns.

Based on the majority vote of multiple indices, *k* = 4 was suggested. However, since one of the four clusters accounted for more than half of the objects (*n* = 25), we decided to proceed with *k* = 5 to explore more fine-grained subtypes. Notably, one cluster included just a single application (“neolexon Aphasie”, an app to support speech therapy), that significantly differentiates from other DiGAs in terms of its characteristics (see below). Figure 3 depicts the dendrogram with the highlighted clusters.

**Figure 3:**
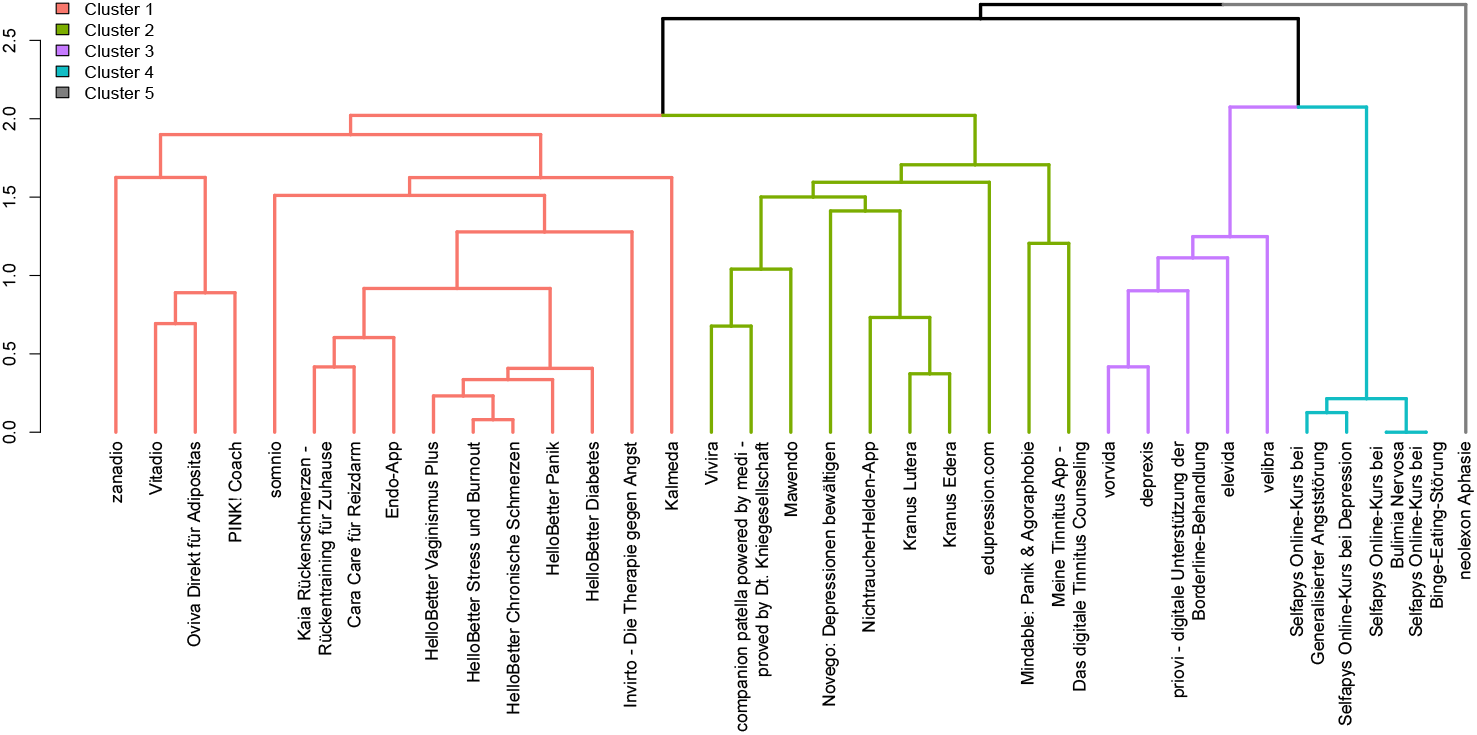
Dendrogram of the first *n* = 35 DiGAs using Ward’s method.

A detailed inspection of the dendrogram reveals that cluster 1 and 2 share a common parent node as well as cluster 3 and 4. Notably, the latter clusters exclusively consist of applications from the same DiGA provider (“GAIA AG” applications in cluster 3 and “Selfapy” applications in cluster 4). Similarly, apps from the provider “HelloBetter” are grouped together, overall suggesting face-validity of the clustering outcomes and reflecting different product strategies. To mitigate potential biases related to the provider’s product portfolio and enhance the generalizability of our results, we decided to merge clusters 3 and 4 for the cross-tabulation analysis. As the fifth cluster only consists of a single DiGA marking a potential outlier, it was excluded from the subsequent analyses because the aim of archetypes is to identify recurring design patterns. However, we discuss the application with its unique feature combinations separately and how it relates to the other archetypes. Table 2 shows the results of the cross-tabulation analysis for all 108 characteristics in the 19 dimensions. Note that the frequency of five characteristics listed in Table 2 is zero across all clusters because only the first-level functional areas were considered, and one DiGA was excluded.

**Table 2:**
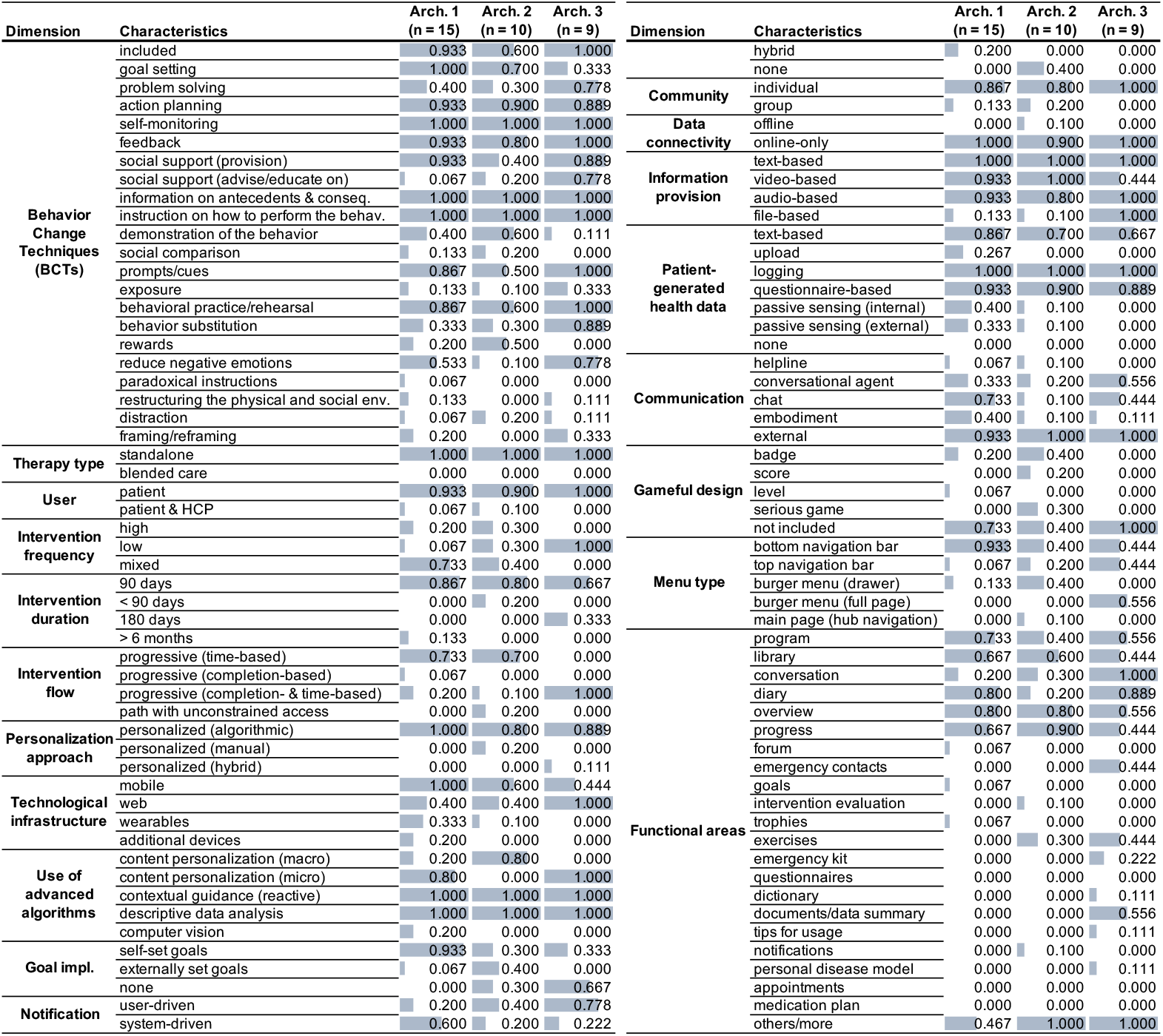
Results of the cross-tabulation analysis (*n* = 34).

We observed a total of 8 characteristics that were integrated in all DiGAs independent of the cluster assignment. These are: i) self-monitoring, ii) information on antecedents and consequences, iii) instruction on how to perform the behavior, iv) contextual guidance (reactive), v) descriptive data analysis, vi) text-based information provision, vii) logging, and viii) a standalone therapeutic approach. Furthermore, this core set of characteristics expands to 20 when accounting for a minimum occurrence of >80% across all DiGAs. In particular, the i) inclusion of BCTs, respectively, explicit grounding in behavioral therapy, ii) action planning, iii) feedback, iv) behavioral practice/rehearsal, v) patient as the only user, vi) no community function, vii) an online-only intervention, viii) algorithmic personalization, ix) the integration of videos and x) audios for providing information, xi) the integration of questionnaires for capturing patient-generated health data, xii) and external communication. In the following paragraphs, the distinguishing characteristics of the identified archetypes are described.

**Archetype 1** is characterized by its strong focus on goal setting (100%), particularly self-set goals (93.3%). Therefore, we denote this archetype as **“personal goal-centered DTx”**. With regard to the technological infrastructure, the intervention is delivered as a mobile app and may be coupled with wearables or additional devices (esp. bluetooth body scale, VR glasses with headphones). This archetype is also characterized by the highest share of system-driven notifications and sensor integration (both internal and external). Sensed parameters of DiGAs in this cluster were daily steps (*n* = 3), body weight (*n* = 2), blood glucose, blood pressure, sleep, burned calories, position of the mobile phone (gyroscope) as well as the position and motion of the user (each *n* = 1). In addition, *n* = 3 DiGAs use the built-in smartphone camera to support nutrition tracking via barcode scanning (*n* = 1) or AI-based food recognition (*n* = 2). Notably, this cluster is also the only one that includes apps using computer vision for context acquisition to promote goal attainment (20%). It is also noticeable that this cluster has the highest share of “chat” as directly integrated communication channel to interact with professional support (73.3%). From a navigation perspective, the UI mainly features a bottom navigation bar (93.3%) and prioritizes direct access to the therapeutic program as the primary functional area (73.3%) compared to the other clusters.

**Archetype 2** offers the largest share of rewards at the content level (BCTs) (50%), reflected through a gameful design with badges and scores at the interface level. Notably, only in this cluster DiGAs can be found that use serious games as a therapeutic element. We thus refer to this archetype as **“gameful designed DTx”**. In addition, this cluster also predominantly consists of apps that personalize the therapeutic program on a macro level and therefore tailor the composition as well as chronological order of the content to the user. Compared to the first archetype, goal setting is also frequently found (70%), however more use is made of externally specified goal values. This suggests that there may be continuum-like transitions between both archetypes. Although some apps (20%) promoted community feelings, social support generally plays a less prominent role compared to the other clusters. Similarly, the diary as a functional area is less prominent, even though insights on individual progress are often available at the first interface level (90%).

Although the single-item clustered DiGA “neolexon Aphasie” also includes several of the mentioned “core-characteristics” of DiGAs, such as self-monitoring and feedback, it clearly distinguishes itself from others by its therapeutic approach, which aligns with the idea of “blended care” by complementing in-person speech therapy. Therefore, the app provides dedicated configuration possibilities for HCPs to prescribe manually tailored exercises and goals for the patient (i.e., external). Furthermore, the intervention focuses solely on skills training, reflected in a very targeted and minimalistic UI structure that centers exercises and settings (others/more) as functional areas at the first level. The app also includes gamification features that allow users to collect stars (badges) when completing the exercises. Therefore, we argue that it most closely may align with this second archetype from a conceptual view.

In **Archetype 3** interventions prominently feature problem solving (77.8 %), social support (provision and also advise on) (88.9%; 77.8%), exposure (33.3%), behavior substitution (88.9%), as well as the reduction of negative emotions (77.8%) at the content level. A distinctive feature of this cluster is its pronounced emphasis on the conversational aspect as a dedicated functional area within the UI (100%). We therefore suggest **“conversational-driven DTx”** as umbrella term for DiGAs of this archetype. This becomes also evident in the highest share of using conversational agents as a communication channel (55.6%). Likewise, chat functions for interacting with a human are represented here second most (44.4%). Additionally, emergency contacts, an emergency kit, a dictionary, and insights from the collected data and documents are centered in the UI. From the perspective of the intervention flow, all applications used a combination of completion- and time-based progression. The integration of notifications is primarily user-driven (77.8%) compared to the other clusters. Furthermore, it is noticeable that no evidence regarding gameful design was found in this cluster (not included).

As mentioned, this cluster consists of *n* = 9 DiGAs which are operated by only two providers. A detailed analysis of the DiGAs belonging to these two branches (cluster 3 and 4 in Figure 3) reveals two different paradigms in human guidance delivery. In particular, DiGAs provided by “Self-apy” employ a “human-guided self-management” approach where a psychologist monitors the patient’s progress and is available via an integrated chat function to ensure patient safety [71]. In contrast, DiGAs provided by GAIA AG aim to simulate this sense of human guidance by encapsulating the entire intervention as a conversation with an agent [72, 73]. This interaction follows a rule-based approach, in which the agent’s responses depend on the patient’s chosen constrained answer options. Compared to other DiGAs with a conversational agent, the dialog is not just an “add-on” alongside other non-dialog-based functions (e.g., program, diary), but rather the heart of the application. Technically, GAIA applications are based on the “broca” platform and therefore follow a very similar approach [74]. Based on these considerations, we propose two sub-archetypes where **3a)** represents **“human-guided self-management”** leveraging computer-mediated communication and **3b)**, a **“simulated therapy dialog”** using conversational agents (as depicted in Figure 4).

**Figure 4:**
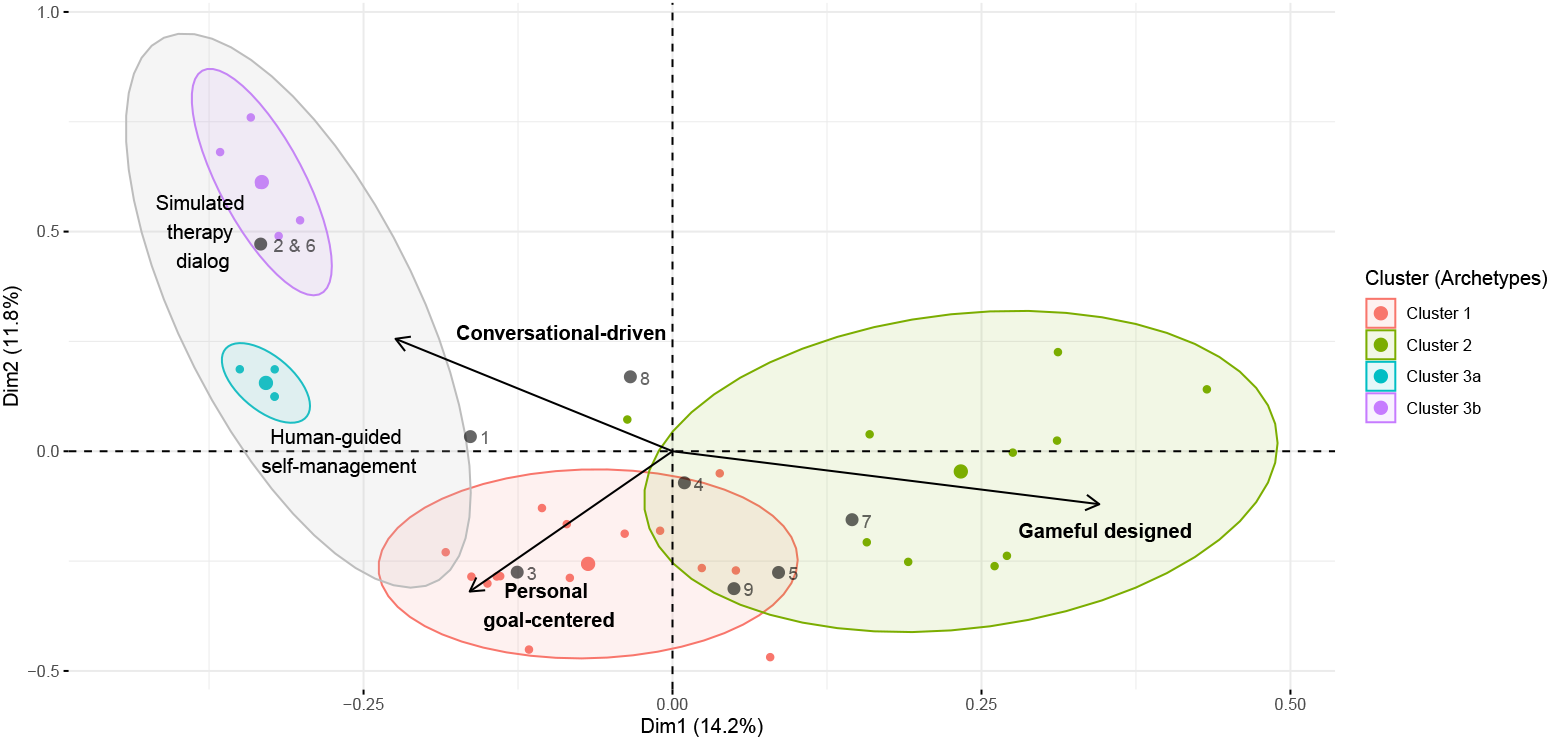
Bi-plot of the first two dimensions of the multiple correspondence analysis (MCA) with ellipses highlighting the clusters (*n* = 34) and mapping of the *n* = 9 additional DiGAs (arrows visualize the direction of the correlation between the two dimensions and the variables self-set goals, inclusion of gamified elements, and conversation).

### 3.4 Evaluation of archetypes

Figure 4 depicts the relationships of the clusters in the two-dimensional space. The defining characteristic of the clusters becomes also graphically evident in Figure 4, when visualizing the direction of the correlation between these variables and the dimensions as arrows (self-set goals, inclusion of gamified elements, and conversation). Furthermore, the visualization reveals overlaps between the archetypes, particularly between personal goal-centered and gameful designed DTx. Note that the DiGA “neolexon Aphasie” was excluded from the plot for visualization purposes as it was a significant outlier in the upper right quadrant. However, the positioning in the upper right quadrant also graphically underscores the proximity to the second archetype (gameful designed DTx). In the following paragraphs, we briefly discuss the classification of the additionally analyzed DiGAs that are numbered and depicted as gray points in Figure 4.

The first point represents the projection of the blended-care DiGA called “elona therapy Depression” (1). Although a direct chat with a professional (e.g., psychologist) or a conversational agent is not integrated, conceptual overlaps exist at the content level with the integration of BCTs like problem solving and advice on social support. In accordance with the other DiGAs belonging to the third archetype, “elona therapy Depression” also focuses on mental health indications. While bidirectional dialog elements are missing, the overall supported therapy approach may still be regarded as “conversational-driven” in a broader sense by aligning in-person psychotherapy sessions with continuous app-based guidance in the patient’s daily life. Since blended care DiGAs are still rare, this might become an additional sub-archetype to expand in the future.

Both the second and sixth DiGA are provided by the GAIA AG (“somnovia” (2), “levidex” (6)) and have been also assigned to sub-archetype 3b indicating face validity of the projection. The third point involves a projection of the DiGA called “HelloBetter Schlafen” (3), which is assigned to the first archetype but is also situated near the third archetype. First, the assignment to the first archetype is valid given the integration of personal goal setting. Second, since DiGAs offered by “HelloBetter” employ a similar approach to “Selfapy” apps, focusing on human-guided self-management interventions with psychologists to improve patient safety [75], the proximity to the third archetype (esp. 3a) appears justified. A distinguishing characteristic at the interface level might be how these conversational functions are integrated (either as a functional area in the first level or at the second navigational level). Overall, this supports the notion that there can certainly be overlaps between archetypes and that these rather tend to have the character of a continuum.

The fourth DiGA called “My7steps App” (4) was projected at the border between the first and second cluster. Similarly to Selfapy and HelloBetter, a human in the loop exists for ensuring patient safety, called “counselors” which however, remain in the background and intervene if dangerous situations are detected (e.g., suicidal intentions). Therefore, the conversational support is arguably even more limited. Although there may be some conceptual overlaps in characteristics among other DiGAs in the first and second clusters, our analysis did not find evidence of gameful design or systematic support for self-set goals, to the best of our knowledge. Based on these considerations, we argue that it may most closely align with the third archetype (3a). Next, the DiGA “Smoke Free - Rauchen aufhören” (5) was projected on the border to the second cluster, which is reasonable since it features badges and small games aimed at distracting patients when they experience the urge to smoke.

The proximity near to the first cluster is also justified given the opportunity to define personal goals. Similarly, the DiGA “NeuroNation MED” (7) is placed in the second cluster, which is also reasonable as it offers cognitive games with badges and scores as a key feature. The DiGA called “Novego: Ängste überwinden” (8) is spatially projected between the second and third cluster, closer to the second. According to the information provided in the DiGA directory, patients have the opportunity to send one message per week about comprehension questions to the Novego team. However, it is also stated that no therapeutic measures will be taken, which may be the reason why the intervention is still considered unguided [76]. Given that interactive games are part of the intervention, the proximity to the second cluster can be justified.

Finally, “ProHerz” (9) is a DiGA integrating external sensing devices for measuring blood pressure, body weight, heart rate and blood oxygenation to support the monitoring of patients with heart failure. Graphically, the point is projected between the first and second cluster. Goal setting is particularly implemented by providing the ability to define target and threshold values for different vital parameters (with HCPs). As mentioned in section 3.3, externally provided goal values are more associated with the second cluster. However, to the best of our knowledge, we found no evidence regarding gameful design in the analysis material. Considering that the provider emphasizes individualization and progress monitoring by patients and HCPs in the DiGA directory, it may be more appropriate to assign it to the first cluster, where other DiGAs using wearables are also placed (e.g., zanadio, somnio).

In summary, the analysis of the additional DiGAs further underscores the applicability of the archetype framework. Although some overlaps naturally exist, we conclude that the identified archetypes support a structured classification and a discussion of new applications.

### 3.5 Network analysis of combination of behavior change techniques

In the following paragraphs, we aim to provide further insights into the combination of BCTs and groups among them using network analysis with community detection employing the Louvain algorithm. Communities can be defined as “…densely connected nodes, with the nodes belonging to different communities being only sparsely connected” [53]. We decided to focus these analyses on the content level (BCTs) as they provide the shared and technology-independent active ingredients of DTx. Furthermore, we intentionally restricted network modeling and community detection to the overall level due to the relatively small size of clusters. This approach aims to produce more stable results regarding the communities and complement the previously described archetypes.

Figure 5 depicts the network analysis of the content level (BCTs) highlighting three communities. In the center of the network, it becomes apparent that the 11 BCTs goal setting (GS), action planning (AP), self-monitoring (SM), feedback (FB) provision of social support (SSP), information on antecedents and consequences (IAC), instruction on how to perform the behavior (IOB), behavioral practice/rehearsal (BP), prompts/cues (P), demonstration of behavior (DB), and the reduction of negative emotions (esp. stress management) (RNE) are frequently combined with each other. The combination of these BCTs can be regarded as the backbone of most DiGAs.

**Figure 5:**
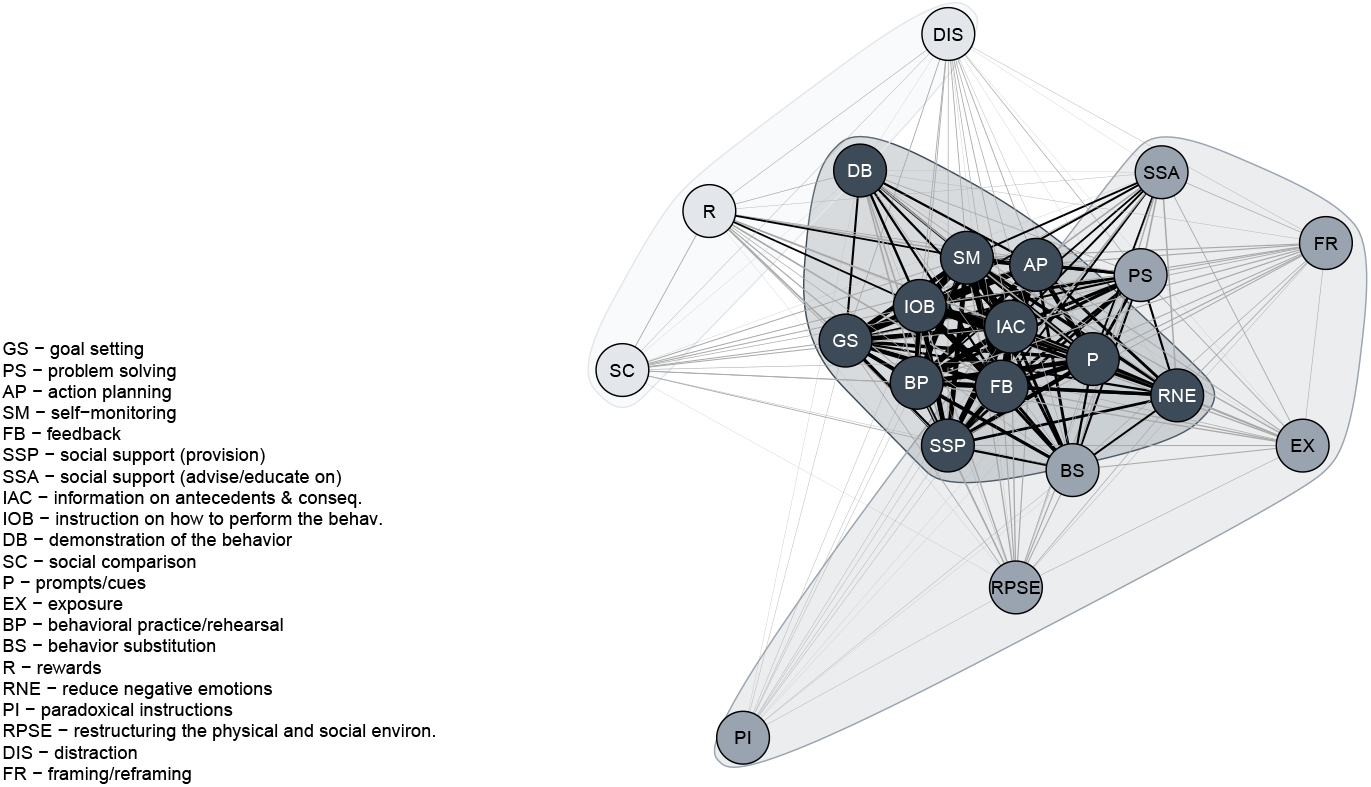
Combinations of BCTs and communities visualized as graph-based network (*n* = 44).

Less frequently, combinations of the BCTs exposure (EX), social support (advise/educate on) (SSA), framing/reframing (FR), paradoxical instructions (PI), restructuring the physical and social environment (RPSE), behavior substitution (BS), as well as problem solving (PS) are found. It is noticeable that behavioral practice is sometimes but not always combined with behavior substitution (including habit reversal) despite belonging to the same cluster of the BCTT [17]. The same applies to problem-solving and social support (advise/educate on) which are also less frequently reported although belonging to the same BCTT cluster like goal setting respectively social support (provision).

Last but not least, the third community consists of distraction (DIS), social comparison (SC) and rewards (R). When rewards are used, they are more often paired with self-monitoring, instructions on how to perform the behavior, and goal setting. This combination is intuitive, as patients know what to do and how to do it, and can monitor their progress toward both the goal and the associated reward. Similarly, when social comparison and distraction are incorporated, they are combined with the aforementioned “backbone” BCTs.

### 3.6 Composition of the user interface of DTx

Despite the growing adoption of digital health interventions, the literature reveals gaps in the systematic examination of the DTx user interface [29]. This is particularly surprising given its role as a critical success factor [77]. While the recent work by Weimar et al. [33] is a first contribution in this regard, a detailed analysis how the interface is composed in terms of navigation and first and second level functional areas, is lacking. To address this gap, we analyzed and modeled the composition of the UI as tripartite graph.

Figure 6 reveals that the most frequently employed approach is the use of a bottom navigation bar. In particular, 61.3% (*n* = 27) of the analyzed DiGAs use this navigation pattern, with on average 4.407 first-level functional areas are accessed via the bottom navigation bar. With descending frequency, the functional areas diary (*n* = 21), progress (*n* = 20), overview (*n* = 19), program (*n* = 18), library (*n* = 14), others/more (*n* = 12), exercises (*n* = 5), conversation (*n* = 3), emergency kit (*n* = 2), forum, goals, notifications, and trophies (each *n* = 1) are navigated from there (first level). Multiple menu type combinations using a bottom navigation bar do exist, but they are relatively uncommon (*n* = 5). Specifically, in the case of *n* = 4 DiGAs (same provider), a combination of a bottom navigation bar and a top navigation bar was observed. Further, *n* = 1 DiGA paired a bottom and top navigation bar with a burger menu (drawer). The analysis of the first and second levels shows that the functional area “overview” also acts as a central navigation component. It presents a dashboard-like view of the subsequent functional areas, also allowing users to directly access conversations, exercises, diary, goals, the program, and progress.

**Figure 6:**
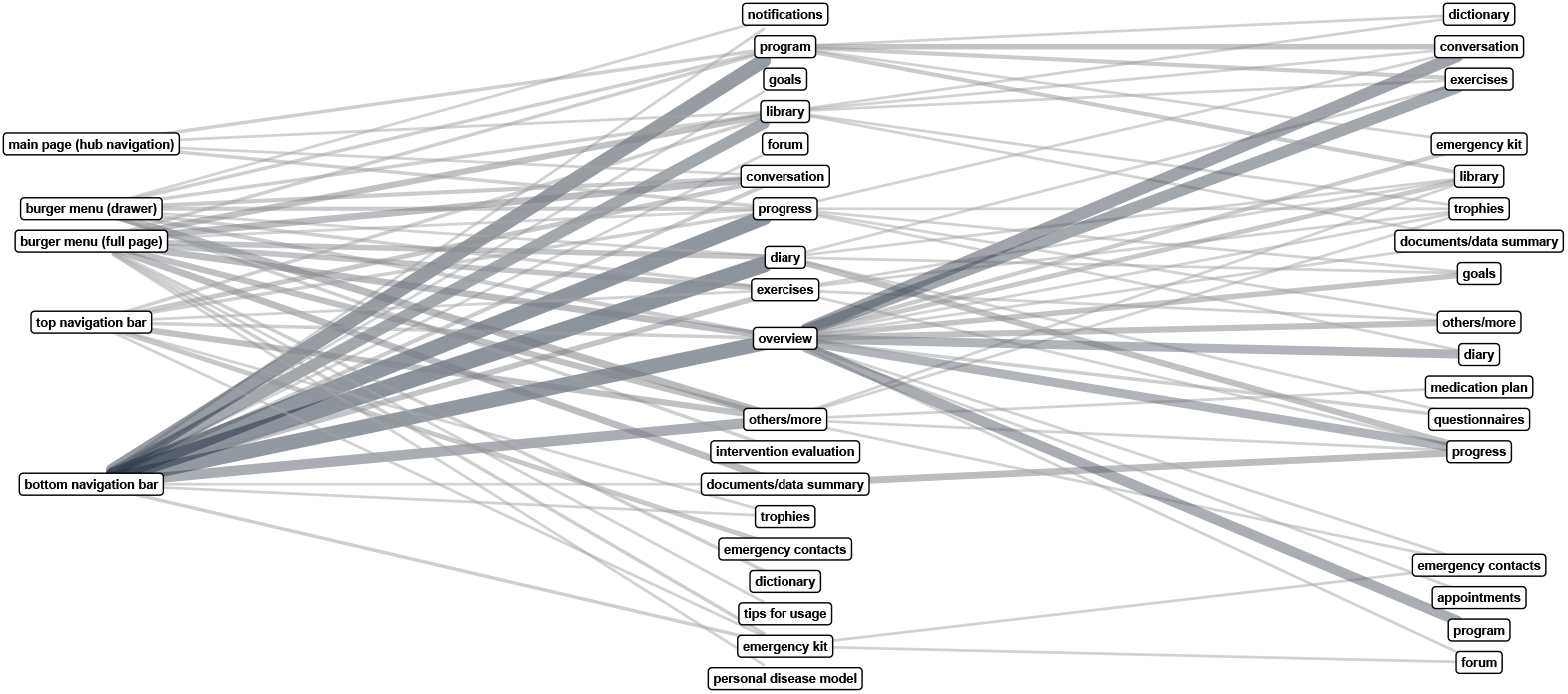
Composition of the user interface modeled as a tripartite graph (*n* = 44).

Beyond bottom navigation, the following other menu types were observed: burger menu (full page) (*n* = 7), burger menu (drawer) (*n* = 6), main page (hub navigation) (*n* = 1) and top navigation bar (*n* = 1).

### 3.7 Exploratory analysis of associations between DTx design and health- mand engagement-related outcomes

By analyzing the approval study design of the *n* = 44 DiGAs, we identified and grouped the primary intervention outcome measures into 9 categories. To guide this categorization and ensure it is not overly detailed given the limited sample size, yet still meaningful for evaluation, we used the outcome domains described in the taxonomy by Dodd et al. [54] as general orientation. Furthermore, given the strong focus of existing DiGAs on mental health (54.5%; *n* = 24), we also complementarily used the hierarchical taxonomy of psychopathology (“HiTOP”) for more depth and guidance in this regard [55].

It is noticeable that the vast majority of studies employed patient-reported outcome measures as primary outcome (i.e., self-reported), where only *n* = 4 DiGAs used “hard” clinical objective outcomes (weight loss, reduction in HbA1c, physical stress test). Because these outcomes stand out from the rest, they have been grouped together in a separate category. Overall, the following nine categories were formed: clinical objective outcomes (*n* = 4), physical symptoms (*n* = 6; non-musculoskeletal), impulsive & dysregulated behavior (*n* = 6; esp. substance abuse, borderline personality disorder, eating pathology), anxiety (and fear) (*n* = 6), depression and related distress (including “burn-out”) (*n* = 9), musculoskeletal outcomes (*n* = 5), physical functioning (sleep & fatigue) (*n* = 4), social, role, and cognitive functioning (*n* = 2), as well as quality of life (*n* = 2). It should be noted that the category “impulsive & dysregulated behavior” diverges from the HiTOP taxonomy by combining externalizing disorders (esp., substance abuse and borderline personality disorder) with disorders related to eating pathology, such as binge eating disorder and bulimia nervosa (internalizing). This decision was based on similarities in impulsiveness and clinical presentation [78].

Figure 7 illustrates the relative co-occurrences of outcome categories and BCTs, indicating that DiGAs aimed at a particular outcome also include these BCTs. Overall, the co-occurrences additionally underscore that the BCTs goal setting, action planning, self-monitoring, feedback, social support (provision), information on antecedents and consequences, instruction on how to perform the behavior, as well as behavioral practice/rehearsal form the core of existing DiGAs independently of the targeted outcomes. The BCT problem solving is found in nearly all DiGAs focusing on improving depression & distress as well as quality of life, but also in those focusing on anxiety and impulsive & dysregulated behavior. Likewise, social support (advise/educate on) was observed particularly in DiGAs focusing on the aforementioned outcome categories. However, some BCTs such as social comparison, exposure, paradoxical instructions, restructuring the physical and social environment, distraction as well as framing/reframing are currently only more selectively used for targeting certain outcome categories. In part, this may be explained by the pathogenesis and therapeutic approach, so it is reasonable that exposure to anxiety-inducing situations is related to anxiety-focused DiGAs. In addition, by restructuring the physical and social environment, the potential triggers of mental disorders can be made more manageable. Similarly, distraction is found in DiGAs targeting impulsive & dysregulated behavior to offer digital alternatives in unwanted situations (e.g., smoking), but also intensifying physical symptoms (e.g., urge to urinate, tinnitus). Notably, rewards have not been identified in our analysis in DiGAs focusing on anxiety, the musculoskeletal system, as well as physical functioning highlighting potential areas for future expansion.

**Figure 7:**
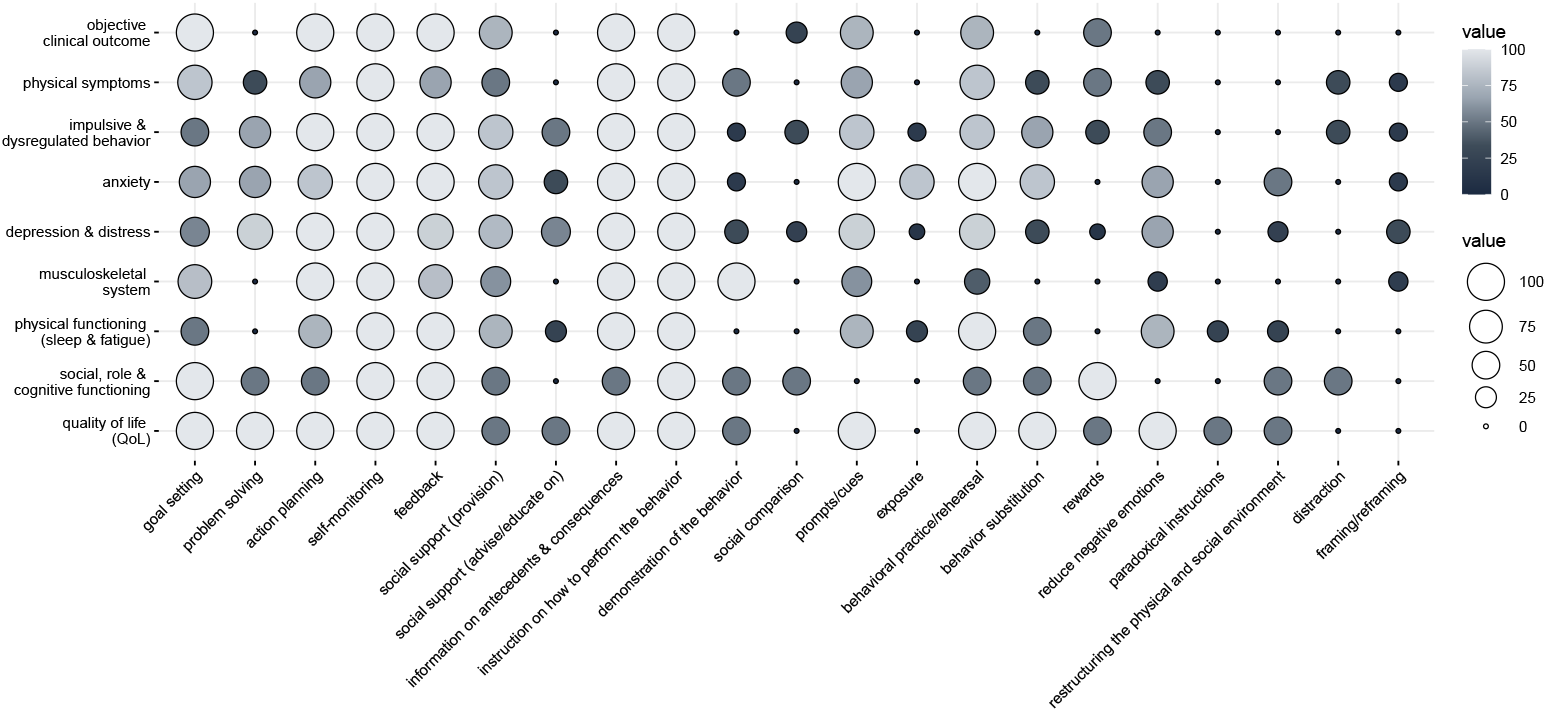
Relative co-occurrences of outcome category and BCTs (*n* = 44).

Beyond the BCTs, we also analyzed the associations between outcome category and the functional areas of the DiGA. For this analysis, the first and second level functional areas were considered in combination. Figure 8 depicts the results of these relative co-occurrences. Correspondingly, as similarly shown in Figure 7, some functional areas have been observed across all of the outcome categories. Specifically, the functional areas library, conversation, diary, overview, progress, exercises, and others/more were associated with all outcome categories. In contrast, the functional areas forum, emergency contacts, goals, intervention evaluation, trophies, emergency kit, questionnaires, dictionary, documents/data summary, tips for usage, personal disease model, appointments and medication plan have been integrated only selectively. This, in turn, opens up opportunities for expanding existing DiGAs, although some imbalances might also be attributed to the varying medical needs associated with different targeted outcomes.

**Figure 8:**
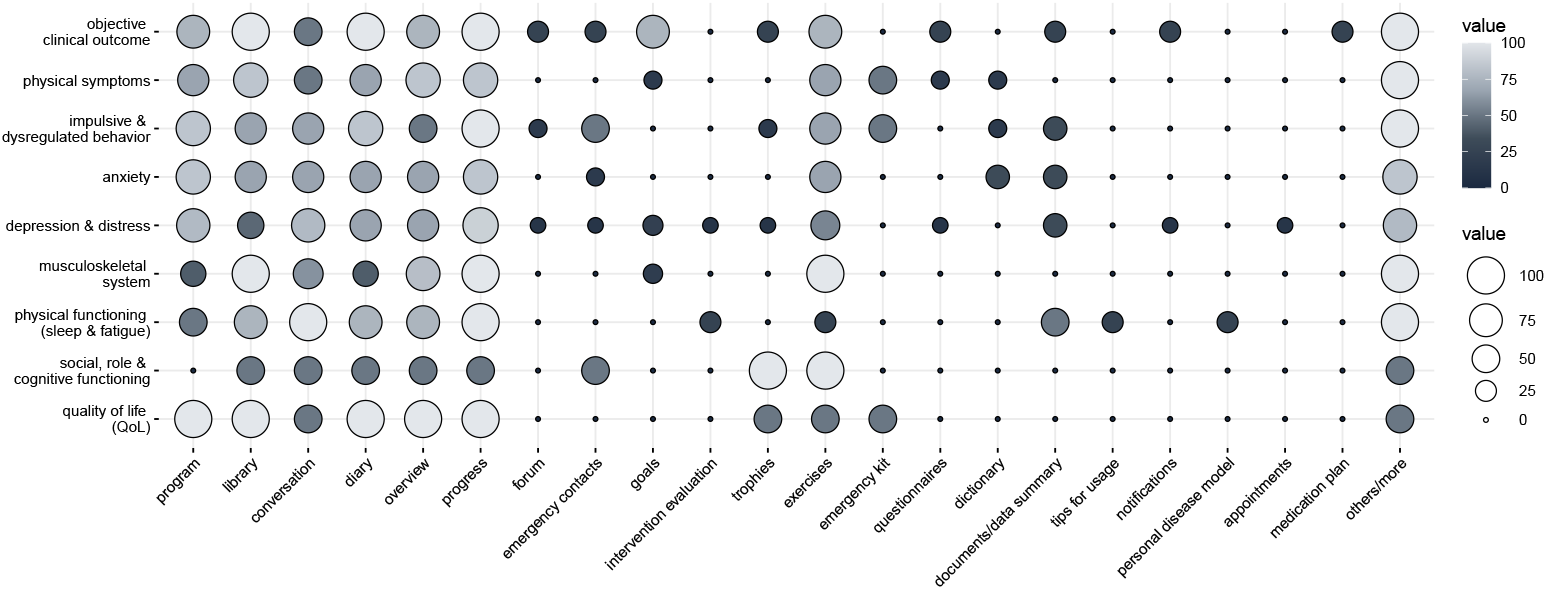
Relative co-occurrences of outcome category and functional areas of the user interface (*n* = 44).

Given that goal setting belongs to the core BCTs of existing DiGAs, it is particularly noticeable that it is scarcely mirrored as a dedicated functional area in the UI. Instead, goals are subsumed in other areas such as progress, diary or program. However, the use of “goals” is comparatively more frequently found in DiGAs targeting “hard” clinical objective outcomes. One possible explanation could be that it is particularly important here to make the goals transparent and easy accessible to the user in order to promote intervention engagement. Likewise, the use of “trophies” as functional area in itself differs from the distribution of rewards shown in Figure 7. Figure 8 also indicates that apps focused on conditions susceptible to sudden changes (esp. unwanted impulses, urges, or unexpected occurrence of symptoms) – such as smoking cessation, alcohol reduction, bladder dysfunction, tinnitus, borderline personality disorder, or irritable bowel syndrome – include a quickly accessible “emergency kit” providing immediate self-management strategies for these situations. Last but not least, our analysis reveals that a DiGA-based support of the patient’s pharmacotherapy (medication plan) has only been found in one app so far. This finding is surprising when one considers that patients can also receive an accompanying medication (e.g., psychotropic drugs) in a variety of scenarios in addition to the DiGA itself. At the same time, this finding underscores that existing DiGAs are still often understood as an “isolated” approach that is less harmonized with other forms of therapy.

Finally, we conducted an exploratory investigation of the taxonomy’s potential as a tool for facilitating data-driven analyses with machine learning techniques. Therefore, we used the app store ratings of the analyzed DiGAs as a proxy for the user engagement and searched for characteristics that best explain the ratings. For this purpose, we fitted three regression trees for each of our meta-characteristics to identify distinguishing characteristics that hypothetically may explain engagement differences. We used the R package rpart [56] which is based on the CART algorithm (classification and regression trees) [79]. Given the limited sample size of *n* = 25 DiGAs with app store ratings, we restricted the maximum depth of the tree to one level.

The first regression tree model (*R*^2^ = 0.071) splits the sample on the BCT “prompts/cues” where the presence of this characteristic was associated with a lower mean predicted rating (4.21) compared to an absence (4.42). Next, a regression tree was fitted on all characteristics related to the delivery logic and technology. This second tree (*R*^2^ = 0.102) splits the data on the characteristic “progressive (time-based)” with the existence predicting a higher mean rating (4.37) than the absence (4.14). Lastly, we fitted a model for the patient’s interface. The regression tree, which considered all interface-related characteristics (*R*^2^ = 0.049), divided the data on whether the UI included a “conversation” element in the first navigational level or not. In particular, the inclusion was associated with a lower predicted rating (4.15) than the absence (4.33).

While these findings should be cautiously interpreted given the limited sample size and the highly simplified assumption of causality, they still shed light on potential differences that could serve as a starting point for the generation of hypotheses. For example, the potential benefit of a time-based progression through the intervention (e.g., unlocking a new module each week) may be explained by a clear and straightforward structure related to time investment that resembles traditional in-person programs [80]. Similarly, prompts/cues operationalized as notifications in existing DiGAs might eventually become burdensome for the patient at a certain dose (frequency), negatively impacting engagement [81]. For conversational elements, one explanation is that they can lead to incorrect user expectations, such as expecting a working alliance similar to face-to-face therapy, which may not be fulfilled [82]. Some of these hypothetical explanations can be partially substantiated by existing studies while simultaneously providing guidance for future research.

## 4 Discussion

The present study systematically developed a comprehensive taxonomy of clinically approved DTx listed in the German DiGA directory, following both a deductive and inductive approach. This allowed abstracting and synthesizing the underpinning elements that form the pillars of all clinically approved DiGAs so far. The various implications of these findings for theory and practice are discussed below.

### 4.1 Theoretical and practical implications

Using the taxonomy, three archetypes are identified underlying current DiGAs. While archetypes have been described for the broad field of mHealth apps in general by Greve et al. [62] or apps for depression by Mueller et al. [66], this work is the first, to the best of our knowledge, that quantitatively clusters the German DTx landscape. Furthermore, going beyond prior research, the present study follows a holistic approach by combining characteristics related to the content, delivery logic and technology, as well as the patient’s interface to extensively understand the design of DTx.

For example, Greve et al. identified five archetypes of mHealth apps where only one of them (“Help me in my Specific Case”) subsumes the concept of DTx. Likewise, the work by Mueller et al. [66] describes the archetype “Certified Internet-based Cognitive Behavioral Therapy Apps” which covers DTx interventions for depression. By describing the three archetypes i) “personal goal-centered DTx”, ii) “gameful designed DTx” and iii) “conversational-driven DTx”, with the latter including two sub-archetypes called a) “human guided self-management” as well as b) “simulated therapy dialog”, this study provides detailed insights into the different forms of DTx and extends previous work. Furthermore, the identified archetypes were validated based on an analysis of *n* = 9 additional DiGAs. Table 3 summarizes the three archetypes and highlights their key characteristics.

**Table 3:**
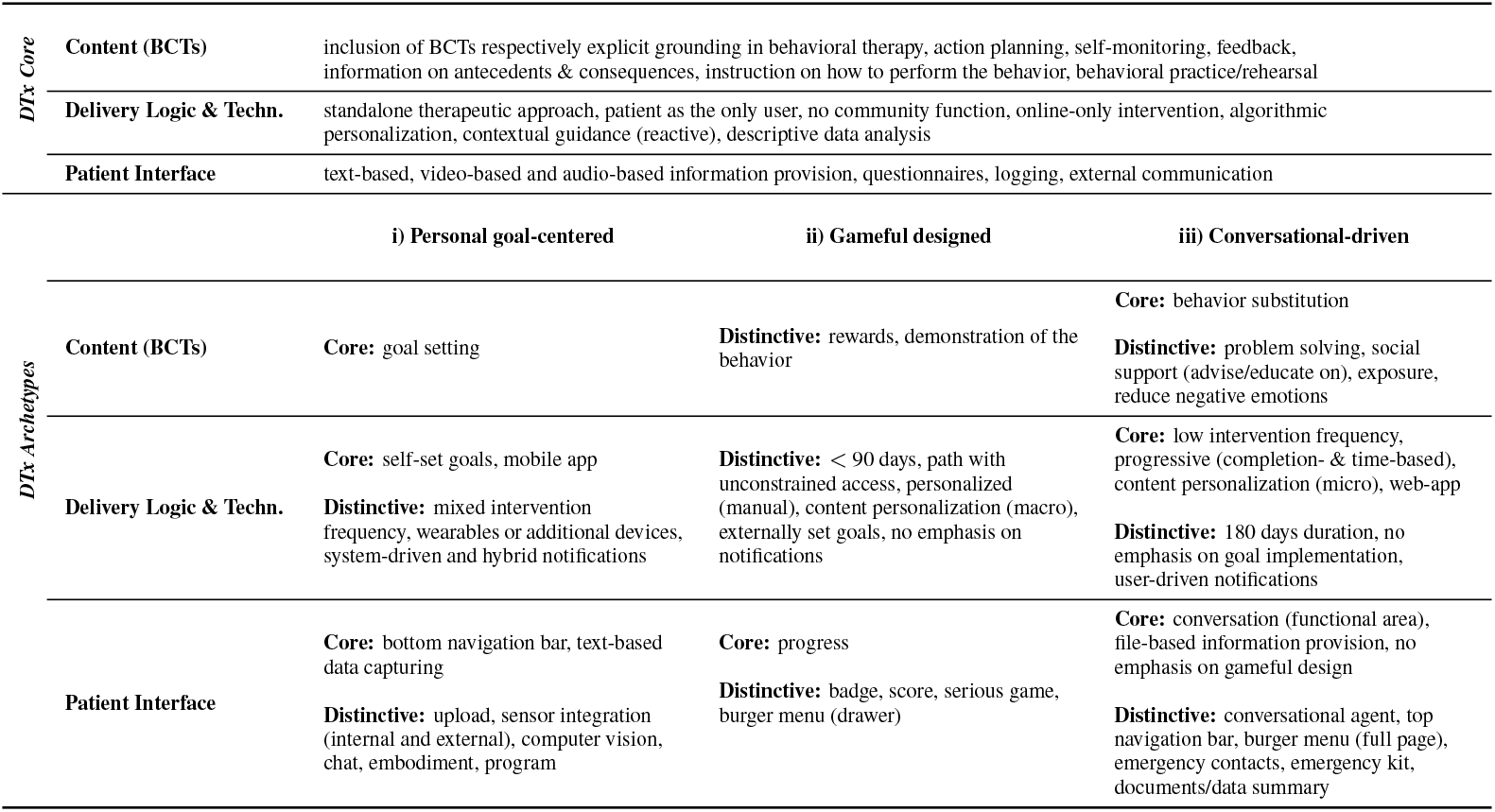
Summary of the overarching characteristics (*>*80% prevalence overall) as well as identified archetypes with core characteristics (*>*80% within a cluster and ≥ 15% higher than in the others) and distinctive characteristics (≤ 80% prevalence but most frequent in the cluster with ≥ 15% difference to the others).

Overall, the derived archetypes enhance the understanding of the contemporary DiGA landscape, which at first glance may appear heterogeneous due to the broad spectrum of medical indications (e.g., mental health, hormones and metabolism, cancer, ears etc.). In addition to classifying existing applications, the archetypes and their relationships (see biplot of MCA in Figure 4) could also inform potential paths for expansion enabling informed decision-making in strategic product management. The positioning within the two-dimensional space could provide a visual framework for identifying incremental and complementary design directions. For instance, when an application currently emphasizes personal goal-centeredness it could be enriched with additional gameful design elements in a next stage of development. Conversely, in the case of a gameful designed app, an approach could be to initially focus on goal-centered aspects before moving on to conversational support. This could ensure that the further development fits to the predominant pattern enabling a coherent DTx design.

Building on the developed taxonomy, additional analyses shed light on the combination of BCTs and the composition of the user interface of DTx. Our findings regarding BCTs align with previous mHealth research with a cross-indication focus, highlighting most of the identified “core BCTs” (e.g., [83, 37]), in particular goal setting, action planning, feed-back, self-monitoring, social support, information on antecedents and consequences, instruction on how to perform the behavior, and prompts/cues. The present study confirms these findings for clinically approved DTx. However, our results may be biased towards mental health indications given the imbalanced distribution in the DiGA directory (e.g., reduce negative emotions).

Although “credible source” was frequently reported as BCT in prior reviews [83, 37], we deliberately did not coded it separately. Our coding relied primarily on provider descriptions, and accurately identifying this BCT would have required examining the entire set of app modules, including specific videos or text passages highlighting experts, which was beyond the scope of this study. Nevertheless, the integration of a conversational element in the interface for interacting with a professional and/or human-like designed agent, may be considered as a possible correlate [37]. When using these characteristics as a proxy, *n* = 31 DiGAs (70.5%) integrated design elements that are associated with the inclusion of a credible source. The true proportion is likely slightly higher, suggesting that credible sources may also serve as a core BCT. Likewise, we were not able to make a clear differentiation between information provision of antecedents and/or consequences, which is why we have considered this as a grouped coding.

While these results shed light on the occurrence of individual BCTs, studies focusing on their combinations (e.g., [84]) are scarce, particularly with a cross-indication focus and for the field of DTx. The present paper contributes to closing this gap by conducting a network-based analysis to uncover the relationships among and between the core and less frequently used BCTs. Beyond the observed core set consisting of eleven BCTs, two additional communities were identified. For example, although the BCT “behavioral practice/rehearsal” is part of the core BCTs, behavior substitution (including habit reversal) was not associated with this core community. This could indicate that current DiGAs might not fully leverage their potential to promote habit formation. Behavior substitution was particularly observed in the conversational-driven archetype suggesting a promising combination. However, to the best of our knowledge, we have found no evidence in the analysis material indicating whether and to what extent advanced habit formation strategies, such as fading reminders [85], are currently used.

While literature recognizes usability as a central barrier regarding the adoption of DiGAs including navigation [86], to the best of our knowledge, no study has yet investigated how the interface of approved DTx interventions is composed. Our analyses reveal that a traditional “bottom navigation” approach with the first level functional areas progress, diary, overview and program was most frequently found. Furthermore, our findings suggest that often multiple access paths to the same functional areas are available, particularly via “overview”. This may be considered as one possible implementation of the usability heuristic “flexibility and efficiency of use” without providing advanced configuration options [87].

To the best of our knowledge, studies that investigated potential associations between targeted health or outcome domains and design-related characteristics are currently lacking. Analyses with a “cross-cutting” focus as in the work by Eaton et al. [83] or Mair et al. [37] are starting to emerge in recent years, where the present study contributes to the scientific discourse in this regard. While the work by Eaton et al. comprehensively evaluated the use of BCTs by the direction of the study results (positive, negative, mixed), it does not dedicatedly analyzes the use of BCTs in dependence to the type of the targeted study outcome. In contrast, the umbrella review by Mair et al. [37] systematically analyzed the inclusion of BCTs in the targeted health domains, claiming “higher-quality evidence from rigorous experimental trials […] to understand which BCTs work for which health domains and in which contexts” as a future research direction. Notably, both studies are limited to the content-level (BCTs). The present paper attempted to close these gaps by i) focusing on clinically approved applications to ensure high quality evidence and ii) broadening the analysis scope to the delivery logic and technology as well as the patient’s interface. This allowed us to identify similarities and differences, which may be partially explained by the nature of the targeted outcomes (e.g., use of an emergency kit area for impulsive or fluctuating disease courses), but also indicate room for further expansion of the existing DiGA landscape. Exploratory regression tree analyses using the app store rating as engagement-related outcome shed additional light on potential discriminating characteristics (prompts/cues, progressive (time-based) and inclusion of conversation as a first level functional area). These analyses should be interpreted cautiously due to the small sample size and *R*^2^ values of 0.1 or less. However, the intervention flow characteristic “progressive (time-based)” showed the highest explanatory power, demonstrating the analytic potential of the taxonomy and suggesting a possible hypothesis for future experimental studies.

### 4.2 Limitations and future research directions

Despite the implications and contributions described above, this study is not without limitations that, in turn, leave room for future research.

First, the scope of our study solely focuses on the German implementation of DTx, namely DiGA. Given that Germany is also a role model for other countries in the field of DTx [21, 10], one could hypothesize that our results may be transferable to other nations. To further substantiate this, future studies could complement this work by building on the proposed taxonomy and repeating the analyses for different countries and markets. A first step towards a cross-country comparison of DTx was recently taken by Liang et al. [88], focusing on high-level technological characteristics (e.g., mHealth, online medical services), targeted diseases, and broad therapeutic functions (e.g., cognitive behavioral therapy, sports therapy). In this regard, our proposed taxonomy could provide a starting point for a more detailed analysis of the “micro-design” perspective of DTx.

In addition, we concentrated on the permanently listed subset of DiGAs that have already shown positive effects on patient care. This group currently makes up the majority of the directory, accounting for 61% as of August 2025. Future research could validate and extend our findings by broadening the scope also to preliminary listed DiGAs and those that have been removed from the directory. Furthermore, in accordance with Weimar et al. [33], the present work only “snapshotted” the DiGA landscape at our time of analysis. Recognizing the dynamic nature of the landscape, our results may, therefore, not remain stable over time. However, they can serve as a reference for “tracking” future development paths. In this regard, future studies could refine the proposed taxonomy and archetypes but also identify potential design-related differences between permanently listed and withdrawn DiGAs.

Second, our analysis material was primarily based on the data provided by the DiGA manufacturers in the BfArM directory (including manuals), in the app stores, and on their linked website. Therefore, it is possible that some features of the analyzed DTx might not be fully mapped in our study. Although two authors conducted the coding and ambiguities in the analysis material were discussed and resolved as thoroughly as possible, misinterpretations and biases cannot be completely ruled out. Consequently, our study is subject to similar limitations regarding the analysis material as described by Weimar et al. [33] whereby, analogously, the combination of various sources was intended to achieve completeness and high accuracy. However, we also examined video demonstrations by “DigaDocs”, scientific and gray literature related to the DiGA, and requested test access whenever there was insufficient material (esp. regarding the UI) for accurate classification. Since we have not tested all DiGAs, the links to the second navigation level of the UI (see Figure 6) must be interpreted with some caution, as it is possible that some links were not captured. Similarly, due to limitations in the level of detail in the analysis material, we were also unable to capture subtle nuances between BCTs related to contexts, actual behaviors or their outcomes. This includes differentiations within broader BCTs such as behavioral practice (e.g., identifying whether context-dependent habit formation as “sub technique” was also targeted), as well as distinctions between BCTs like self-monitoring, feedback, and goal-setting (e.g., setting of outcome and/or behavior goals) [17]. Nevertheless, we argue that our analyses still allow us to uncover the fundamental patterns.

Third, there may be a bias in our results towards mental health apps as they represent a large part of the analysis set (a total of 24 from 44; 54.5%). The third archetype (conversational-driven) largely consists of mental health DiGAs, with one exception in the additionally analyzed DiGAs. Nevertheless, the analysis shows that mental health applications were still implemented differently and can be found in all clusters. This highlights the importance of systematically examining DTx design across various dimensions, as demonstrated in this work.

While some possible directions for future research have already been mentioned, there are others as well. For example, given the three identified “high-level” archetypes, future research may investigate how they can be effectively combined into one artifact. This may include whether the combination has downsides (e.g., technostress) or benefits (e.g., improved engagement and health outcomes). Similarly, some design features have been rarely used so far, but could be potentially advantageous for a variety of apps. For example, only one DiGA was identified that employs an embodied conversational agent (ECA) acting as a visually animated human-like coach (called Albert in “somnio” [89]). Although ECAs may better resemble a face-to-face therapy session than purely text-based chatbots, there is limited evidence about using them in professional care settings [90, 91]. Specifically, how they should be designed and which tasks and target groups they are best suited for remain areas of ongoing research [92]. Combined with the capabilities of large language models (LLMs), ECAs could achieve a higher level of sophistication and open up new opportunities for DTx enhancement. Furthermore, recent research emphasizes the potential of LLMs to enable highly personalized “just-in-time adaptive interventions” (JITAIs), providing proactive support to patients precisely when it is most needed, filling a gap in the current DiGA landscape. [93]. Although “LLM-driven DTx” serving as longitudinal health companions are currently non-approvable under existing regulatory frameworks, Freyer et al. suggest they might be approved in the future with “minor to medium changes” in regulations [94]. Building on the concept of multimodal LLMs, various patient inputs like audios, images (such as medical images from electronic health records), videos (e.g., for guiding exercises) and text can be combined to personalize the intervention [95]. However, ensuring patient safety is essential, and appropriate strategies are necessary to disseminate the potential of LLMs in medical care [96].

Last but not least, our results also provide a further step towards “Meta-DTx” as proposed by Fürstenau et al. [23]. These integrate and align DTx for multimorbid patients in one single application. The proposed taxonomy could guide the technical design for such orchestrator apps and guide the refinement of existing data exchange standards. Finally, our taxonomy could accelerate the development of no-code authoring tools supporting lay developers (e.g., physicians or psychologists) to create and adapt DTx applications without programming knowledge [97, 98]. LLMs could also play a role in this regard. In summary, this could reduce development costs and time to deliver benefits to patients, while also enabling new care models like blended care.

## 5 Concluding remarks

With the rise of digital therapeutics (DTx) as patient-facing systems, digital opportunities exist to continuously support patients and promote adherence in daily life. The current study proposed a new taxonomy for classifying DTx, considering both technical and behavior change aspects. A clustering analysis was used to uncover three archetypes in the examined German DTx landscape (DiGAs). The results revealed the integration of a core set of behavior change techniques (BCTs) in the vast majority of DiGAs. Our additional findings tackled the gap in systematically examining how BCTs are combined, how the patient’s interface is designed, and how design characteristics relate to different health and engagement outcomes. Focusing on these underexplored areas within a sample of clinically approved applications, our study offers new insights with theoretical and practical implications for the field of DTx.

## Acknowledgements

During the preparation of this work, the authors used Grammarly and Chat-GPT (4o, 5 & 5.1) to correct typographical errors and enhance grammar and clarity. After using these tools/services, the authors reviewed and edited the content as needed and take full responsibility for the content of the published article.

## Ethical considerations

This study used publicly available information and did not require ethical approval since no primary data from human participants was collected.

## Consent to participate

Informed consent was not required for this study as we used publicly available information and no data from human participants was collected.

## Consent for publication

All authors have provided their consent.

## Funding

This study did not receive any funding.

## Declaration of conflicts of interest

The authors declared no potential conflicts of interest with respect to the research, authorship, and/or publication of this article.

## Data availability

The data that support the findings of this study are available from the corresponding author upon reasonable request.

## A Description of the taxonomy dimensions and characteristics

The Tables 4, 5 and 6 summarize all dimensions and characteristics along with definitions of DiGATax to support accurate coding. Please take note of the following points:

1. Alignment means that the conceptualization used in DiGATax at least includes the aligned concept(s) (i.e., is comparable or broader than the referenced concept(s)). Likewise, when a previous taxonomy proposed several closely related characteristics within the same work, these may have been mapped to a single, broader characteristic in DiGATax. However, in the case of dimensions only a 1:1 mapping was applied.
2. A referenced dimension or characteristic was mapped only once in the taxonomy within this work, to the most appropriate position, unless the characteristic (including its definition if available) combines multiple concepts, such as “audio and video”, which are represented separately in DiGATax. In this case, they are split (e.g., “audio-based” and “video-based”).
3. “Aligns with” on the level of a dimension means that a corresponding dimension was found in the referenced work (with or without overlapping characteristics).
4. “Aligns with” on the level of a characteristic means that one or more corresponding characteristic(s) were found (may have been assigned to a different dimension in the referenced work and vice versa).
5. If there were significant terminological deviations despite conceptual overlap, the related terms were also named.
6. If definitions were adopted this was indicated as “adopted from” and (if available) a reference to the numbering code in the referenced work.

**Table 4:** Description of taxonomy dimensions and characteristics related to the content.

*\* = non-mutually exclusive dimensions*
| No. | Code | Definition | Aligns with |
| --- | --- | --- | --- |
| 1 | **Behavior Change Techniques (BCTs)\*** | BCTs are the “active ingredients” [17] of a behavioral intervention. | [65] |
| 1.1 | included | The provider explicitly reports that BCTs are incorporated and/or the grounding in (cognitive) behavioral therapy. Note: even if the application is based on a different therapeutic approach (e.g., exercise therapy, speech therapy), BCT analogs may still be identified. In this case, included is not coded, but the identified BCT analogs are coded accordingly. |  |
| 1.2 | goal setting | adopted from BCTT [17] 1.1, 1.3 |  |
| 1.3 | problem solving | adopted from BCTT [17] 1.2 |  |
| 1.4 | action planning | adopted from BCTT [17] 1.4 | [15] |
| 1.5 | self-monitoring | adopted from BCTT [17] 2.1, 2.3, 2.4, 2.5, 5.4 | [15] |
| 1.6 | feedback | adopted from BCTT [17] 2.2, 2.6, 2.7 |  |
| 1.7 | social support (provision) | adopted from BCTT [17] cluster 3<br>Note: this characteristic focuses on the direct provision of social support. |  |
| 1.8 | social support (advise/educate on) | adopted from BCTT [17] cluster 3<br>Note: this characteristic refers to the indirect way of providing social support by advising or educating the user on how to receive support from others. |  |
| 1.9 | information on antecedents & consequences | adopted from BCTT [17] 4.2, 5.1, 5.3, 5.6 | [15] |
| 1.10 | instruction on how to perform a behavior | adopted from BCTT [17] 4.1 |  |
| 1.11 | demonstration of the behavior | adopted from BCTT [17] 6.1 |  |
| 1.12 | social comparison | adopted from BCTT [17] 6.2 |  |
| 1.13 | prompts/cues | adopted from BCTT [17] 7.1 | priming, just-in-time reminders [15] |
| 1.14 | exposure | adopted from BCTT [17] 7.7 |  |
| 1.15 | behavioral practice/rehearsal | adopted from BCTT [17] 8.1.<br>Note: this may include habit formation [8.3] if the context association is also targeted. | train self-control [15] |
| 1.16 | behavior substitution | adopted from BCTT [17] 8.2<br>Note: this may include habit reversal [8.4] if rehearsal and repetition are emphasized. | train context-response [15] |
| 1.17 | rewards | adopted from BCTT [17] cluster 10 | revalue outcome [15] |
| 1.18 | reduce negative emotions | adopted from BCTT [17] 11.2 |  |
| 1.19 | paradoxical instructions | adopted from BCTT [17] 11.4 |  |
| 1.20 | restructuring the physical and social environment | adopted from BCTT [17] 12.1, 12.2. | alter context, alter cue salience [15] |
| 1.21 | distraction | adopted from BCTT [17] 12.4 |  |
| 1.22 | framing/reframing | adopted from BCTT [17] 13.2 |  |

**Table 5:** Description of taxonomy dimensions and characteristics related to the intervention delivery logic and technologies.

*\* = non-mutually exclusive dimensions*
| No. | Code | Definition | Aligns with |
| --- | --- | --- | --- |
| **2** | **Therapy type** | Specification of the extent to which the application supports in-person therapy (face-to-face). | [66], route [67] |
| 2.1 | standalone | No additional support by an HCP is required, i.e., only patient-oriented intervention. | [66] |
| 2.2 | blended care | Intervention must be accompanied by an HCP, i.e., a dedicated HCP interface is contained or at least the in-app option to adapt the intervention to the patient. |  |
| **3** | **User** | Perception of the patient's role within the intervention. | [62, 31] |
| 3.1 | patient | The patient is the user of the intervention. | [62, 31] |
| 3.2 | patient & HCP | Both the patient and the prescribing HCP can access the intervention. | [62, 31] |
| **4** | **Intervention frequency** | Recommendation by the DTx provider how often patients should interact with the application. Note: these recommendations could differ between functional areas of the DiGA. For example, the diary should be used daily, while the educational modules are intended to be used once a week. In this case, mixed is coded. | [67] |
| 4.1 | high | It is recommended to interact daily, ranging from once to several times a day. | [67] |
| 4.2 | low | Interaction is recommended less often than daily, such as once or a few times a week (or less frequently, e.g., bi-weekly). | [67] |
| 4.3 | mixed | Some functional areas require a more frequent interaction than others. |  |
| **5** | **Intervention duration** | Recommended prescription period by the DTx provider. Note: all DiGAs have a prescription period of 90 days; if no additional information is provided, this default is used as the recommended intervention duration. | [67] |
| 5.1 | 90 days | The use for 90 days (3 months) is recommended for a positive impact. | [67] |
| 5.2 | < 90 days | The use for at least x days (less than 90 days) is recommended for a positive impact and/or the entire intervention program is scheduled for less than 90 days. |  |
| 5.3 | 180 days | The use for 180 days (6 months) is recommended for a positive impact. | [67] |
| 5.4 | > 6 months | Use for more than 6 months is recommended for a positive long term impact. |  |
| **6** | **Intervention flow** | Temporal mode of therapy delivery and determination of intervention content. |  |
| 6.1 | progressive (time-based) | There is evidence for a time-based progression through the intervention program (e.g., every week a new module is unlocked). |  |
| 6.2 | progressive (completion-based) | There is evidence for a completion-based progression through the intervention program (e.g., a new module is unlocked when the previous one was completed). |  |
| 6.3 | progressive (completion- & time-based) | Combination of time-based and completion-based progress. |  |
| 6.4 | path with unconstrained access | Intervention content is organized in a path-like structure with the possibility for flexible on demand access (sequential or non-sequential). |  |
| **7** | **Personalization approach** | Describes the leading actor in personalizing the intervention flow. | [66] |
| 7.1 | personalized (algorithmic) | The system automatically personalizes the intervention flow through an algorithm. Note: some human input may still be necessary. However, the details of the exact outcome of the personalization and the structure of the algorithm are hidden from the user. | [66] |
| 7.2 | personalized (manual) | The intervention flow is explicitly personalized by a human instance (e.g., patient or HCP). |  |
| 7.3 | personalized (hybrid) | Both algorithmic and manual personalization approaches are implemented. |  |
| **8** | **Technological infrastructure\*** | Required technologies for intervention delivery. | [60, 31, 66, 67] |
| 8.1 | mobile | The intervention is accessible through a mobile application. | [60, 31, 66, 67] |
| 8.2 | web | The intervention is accessible through a web-application. | [31, 66] |
| 8.3 | wearables | The application integrates wearables in the intervention to acquire health-related data. Note: this also covers health apps for data synchronization that directly interface with wearables (e.g., Apple Health, Google Fit). | [60, 31, 64] |
| 8.4 | additional devices | The application integrates additional devices or hardware for the treatment, e.g., augmented or virtual reality glasses (AR/VR). | [60] |

Table 5: Description of taxonomy dimensions and characteristics related to the intervention delivery logic and technologies. (continued) *\* = non-mutually exclusive dimensions*
|  |  |  |  |
| --- | --- | --- | --- |
| **9** | **Use of advanced algorithms\*** | Purpose of more advanced algorithms in the intervention. |  |
| 9.1 | content personalization (macro) | Tailoring of the composition and chronological order of content (“macro-level” / “inter-level”). This can, for example, be achieved through an initial profiling questionnaire (e.g., demographic background, preferences) that “maps” the user to a specific path or through ongoing feedback. |  |
| 9.2 | content personalization (micro) | Tailoring within content blocks that are the same for multiple users (“micro-level” / “intra-level”). An example is a branching logic within a psychoeducational module that covers the same topic for multiple users (e.g., coping with relapse situations) but tailors the exact content based on each individual’s interaction (e.g., patient’s prior responses to certain questions). |  |
| 9.3 | contextual guidance (reactive) | Algorithms are used to provide direct feedback during interaction. This includes warnings, recommendations, the deactivation of modules, or adaptation of therapy when the patient cannot follow due to changes in the health condition. | [61] |
| 9.4 | descriptive data analysis | Algorithms are used to process the patient’s health data in a descriptive manner. This could also include visualization, e.g., through statistics or graphs and data aggregation such as averaged values. Note: this can be seen as a technical implementation of self-monitoring. |  |
| 9.5 | computer vision | Algorithms are used to understand visual context, e.g., ingredient extraction in meal images or motion capturing during exercises. |  |
| **10** | **Goal implementation** | Technical correlate of the BCT goal setting. | [69] |
| 10.1 | self-set goals | The user can set goals entirely on their own or choose from predefined ones. | [69] |
| 10.2 | externally set goals | Goals are determined by an external source, e.g., the attending HCP or the digital intervention. | [69] |
| 10.3 | none | No evidence for the integration of goal setting is found in the used information sources. |  |
| **11** | **Notification** | Notification to the user via a dedicated communication channel (e.g., push notification via app, e-mail, SMS), including alerts. | [62, 63] |
| 11.1 | user-driven | Notifications set by the user, e.g., self-timed reminders for mood tracking or medication intake. | [61, 64, 66] |
| 11.2 | system-driven | System initiated notifications, e.g., summaries, motivation, in-app alerts, system-timed reminders, automated messages. | [61, 63, 59] |
| 11.3 | hybrid | Both types of notifications are integrated. |  |
| 11.4 | none | No evidence for the integration of notifications, e.g., for reminders, found in the used information sources. |  |
| **12** | **Community** | Perception of the patient’s role within the intervention. | [61, 62] |
| 12.1 | individual | The focus of the intervention relies solely on the individual user. | [62] |
| 12.2 | group | The intervention emphasizes the community aspect by providing means to help users feel more connected to others (e.g., through an online forum for interacting with peers). | [61, 62] |
| **13** | **Data connectivity** | Possibility to use intervention features without a stable internet connection. Note: online use is default, therefore the categorization is done for online-only and offline. | [62] |
| 13.1 | offline | The intervention can be also used without an internet connection. | [62] |
| 13.2 | online-only | The intervention requires an internet connection and cannot be used offline. | [62] |

**Table 6:** Description of taxonomy dimensions and characteristics related to the patient interface.

*\* = non-mutually exclusive dimensions*
| No. | Code | Definition | Aligns with |
| --- | --- | --- | --- |
| **14** | **Information provision\*** | Means/modalities of technical information provision (delivery) to the user. Note: image-based information provision is not coded separately, as the use of icons and illustrations is self-evident and present throughout all analyzed applications. | [31, 66] |
| 14.1 | text-based | Information is provided via text. | [61, 31, 66] |
| 14.2 | video-based | Information is provided through videos (including animations). | [61, 31, 66] |
| 14.3 | audio-based | Information is provided via audio. | [61, 66] |
| 14.4 | file-based | Information is provided through external sources, e.g., PDFs, sent via e-mail or possible to download. |  |

Table 6: Description of taxonomy dimensions and characteristics related to the patient interface. (*continued*) *\* = non-mutually exclusive dimensions*
|  |  |  |  |
| --- | --- | --- | --- |
| **15** | **Patient-generated health data\*** | Means of technical realizations for information provision from the patient to the system to capture data (including patient-reported outcomes). | [61, 62, 63, 64, 66, 67, 33] |
| 15.1 | text-based | The user can add data in text form, e.g., in a digital diary. | [33] |
| 15.2 | upload | The user can upload visual data, such as images, e.g., to capture a meal, as well as videos or documents. |  |
| 15.3 | logging | Tracking of certain information, e.g., through a scale or a checklist for water intake. This is more user-driven. |  |
| 15.4 | questionnaire-based | Tracking of certain information through a (standardized) questionnaire (e.g., PHQ-9 for reporting severity of depression). This is more system-driven. | [33] |
| 15.5 | passive sensing (internal) | Patient data is recorded using directly connected internal sensors of the device, e.g., use of the smartphone camera (for pose estimation, motion capturing, or nutrition tracking etc.) or integrated step counter. | [62, 63, 64, 33] |
| 15.6 | passive sensing (external) | Data is captured with external devices such as a sensor for continuous glucose monitoring or external synchronized apps (such as Apple Health or Google Fit) and wearables (e.g., smartwatches). | [62, 63, 64, 33] |
| 15.7 | none | No patient-generated data is intended to be captured as an essential intervention component. Note: if a user is solely asked to insert data within an exercise (e.g., fill-in-the-blank text as part of speech therapy), it is also categorized here. |  |
| **16** | **Communication\*** | Technical modes to facilitate communication between the patient (user) and humans or human-like software agents for providing support (or evoke the feeling of being supported). | [61, 62, 63, 66, 67] |
| 16.1 | helpline | Immediate support on symptom worsening or the occurrence of unusual symptoms. | [61, 64, 66, 59] |
| 16.2 | conversational agent | The intervention integrates a conversational agent (e.g., chatbot) that enables constrained and/or unconstrained interaction in natural language. This type of communication is considered synchronous. | [61, 64, 59] |
| 16.3 | chat | Communication with a HCP, treatment companion(s), or technical support is done via chat (primarily text-based) that is directly integrated in the application. Depending on the concrete implementation and responding behavior, this can be more asynchronous or synchronous. Note: this characteristic also includes forums for chatting. | [61, 63, 64] |
| 16.4 | embodiment | The intervention intentionally uses one or more personas with a visual representation (video/animated or static) that embody social roles and accompany the patients throughout the program. If a persona serves as an embodiment of a conversational agent only 16.2 is coded. |  |
| 16.5 | external | Communication with a treatment companion (e.g., HCP) or technical support is not directly part of the intervention, e.g., requests via e-mail or telephone, or through FAQs. | [61, 64, 66] |
| **17** | **Gameful design\*** | Integration of game-like elements as part of the intervention. |  |
| 17.1 | badge | Patients receive a badge when achieving a certain milestone or goal within the intervention. |  |
| 17.2 | score | Patients can collect points, e.g., by solving tasks or completing exercises and modules. | [61] |
| 17.3 | level | Patients can move to a higher level once they successfully complete the lower levels. While this can be in line with a completion-based intervention flow, the gameful design should also be evident in the terminology used within the application. | [61] |
| 17.4 | serious game | Serious games are part of the intervention. |  |
| 17.5 | not included | Gameful design elements are not included. |  |
| **18** | **Menu type of UI\*** | The way the user interface (UI) components are arranged and navigated. | [60] |
| 18.1 | bottom navigation bar | Navigation elements are positioned in a row along the bottom of the UI. |  |
| 18.2 | top navigation bar | Navigation elements are positioned in a row along the top of the UI. |  |
| 18.3 | burger menu (drawer) | Navigation elements are located in a drawer that can slide out (horizontally or vertically) when clicking the menu button. |  |
| 18.4 | burger menu (full page) | Navigation elements are located in an overlay page that opens when clicking the menu button. |  |
| 18.5 | main page (hub navigation) | Navigation elements are centrally located on a main page (hub) from which the different functional areas can be accessed. |  |

Table 6: Description of taxonomy dimensions and characteristics related to the patient interface. (*continued*) *\* = non-mutually exclusive dimensions*
| No. | Code | Definition | Aligns with |
| --- | --- | --- | --- |
| **19** | **Functional areas\*** | Functional areas concentrating the app's capabilities and that are accessible through the user interface. Note: one functional area could be an entry point to navigate to an other area. |  |
| 19.1 | program | Functional area focusing on knowledge building and training of practical skills with a more system-determined intervention flow (structured guidance). This could also include testing of the acquired knowledge using quizzes. Note: when knowledge and skills training are integrated and conducted immediately via a dialog, it is coded as "conversation". If the functional area's focus is only on skills training related to the execution of tasks by the patient, then it is coded as "exercises" instead of "program". | [61, 63, 59, 33] |
| 19.2 | library | Functional area enabling on-demand access by the user to resources for knowledge and skill development (e.g., audios, videos, articles, recipes, working sheets). If the focus is purely on an on-demand accessible collection of tasks to be conducted by the patient or working sheets, "exercises" is coded. | [63, 59, 33] |
| 19.3 | conversation | Functional area that enables a dialog (e.g., with HCP, customer support, chatbot). The dialog may also include active ingredients such as feedback, information provision (educational), or social support. | [33] |
| 19.4 | diary | Functional area that allows patients to self-report their data and may also provide access to historical entries, including data visualization. Note: self-reporting and visualization could be also decoupled as separate functional areas. In this case, "diary" and "progress" are coded. | [61, 64, 59] |
| 19.5 | overview | Functional area that consolidates information about upcoming steps (e.g., planned exercises, educational modules, questionnaire/diary entry completion etc.), the patient's current status, and direction to the specific areas in a dashboard-like manner. Note: this area is also often called "home". | [61, 33] |
| 19.6 | progress | Functional area that provides statistical insights which may be presented as a chart and/or table. This may also include aggregated data (e.g., average) and filtering options (e.g., last week, last month). | [61, 33] |
| 19.7 | forum | Functional area that enables exchange with other users, particularly peers. | [61, 59, 33] |
| 19.8 | emergency contacts | Functional area that enables fast access to emergency hotlines (e.g., in the case of suicidal intentions), telephone counseling or other points of contact (e.g., HCPs on behalf of the provider, counseling centers). If the displayed information includes only the DTx provider's email and/or postal address, this characteristic is not coded. |  |
| 19.9 | goals | Functional area that enables goal setting and/or provides structured access to intervention goals. This may also include presentation of progress toward achieving the goals. In the case goal setting and progress are decoupled, both "goals" and "progress" are coded. | [61, 33] |
| 19.10 | intervention evaluation | Functional area that allows users to rate and give feedback on the overall DTx intervention (e.g., helpfulness, attractiveness of design). |  |
| 19.11 | trophies | Functional area offering access to the collection of symbolic rewards accumulated during the intervention (esp. badges, scores achieved). | [33] |
| 19.12 | exercises | Functional area supporting the conductance and/or management (e.g., planning) of tasks. Note: this can be more system-driven (e.g., by instructing physical exercises), more user-driven (e.g., list of tasks that the patient can mark as completed, downloading of exercise material), or a combination of both. If all exercises have the goal to support the patient in acute episodes associated with an increase in symptoms such as unwanted behavior, "emergency kit" is coded instead. | [61, 33] |
| 19.13 | emergency kit | Functional area that encapsulates material and dedicated exercises (e.g., problem solving strategies, small games, listen to sounds) that aim to guide the patient in acute episodes associated with an increase in symptoms, including unwanted behavior (e.g., tinnitus, sudden mood swings, urge to smoke, drink or urinate). | [66] |
| 19.14 | questionnaires | Functional area that enables the completion of structured questionnaires. This may also include the retrieval of previously completed questionnaires. Note: this characteristic is only coded if the questionnaires are not directly part of a diary. |  |
| 19.15 | dictionary | Functional area that provides explanations of important terms related to the disease and the treatment. |  |
| 19.16 | documents/data summary | The functional area provides access to all documents that have been activated for the patient during the application usage (such as PDF files of information or worksheets), documents uploaded by the patient (e.g., prior medical reports), and data entered into the system (e.g., questionnaires). | [61, 63, 59, 33] |
| 19.17 | tips for usage | Functional area that provides guidance regarding the optimal use of the intervention. | [33] |
| 19.18 | notifications | Functional area that provides access to the notifications sent by the system and/or other users, such as HCPs. Note: this is purely unidirectional, while "conversation" is inherently bidirectional. | [33] |
| 19.19 | personal disease model | Functional area offering access to a personalized disease model developed with the patient during the intervention, aimed at enhancing understanding of how contextual factors interact and influence the disease progression. |  |
| 19.20 | appointments | Functional area to display and manage in-person appointments. Note: in the applications analyzed, this area was used solely for "offline" self-management and was not synchronized with the HCP's schedule. |  |
| 19.21 | medication plan | Functional area to display and manage the medication plan, including the documentation of the medication intake and reminders. | [64] |
| 19.22 | others/more | Functional area referring to multiple administrative functions such as user profile (e.g., name, age, diagnoses) and settings (e.g., password, connection to sensors). | [33] |

## Notes

### Competing Interest Statement

The authors have declared no competing interest.

## References

[1] S. Kraus, F. Schiavone, A. Pluzhnikova, and A. C. Invernizzi. “Digital transformation in healthcare: Analyzing the current state-of-research”. In: Journal of Business Research 123 (Feb. 2021), pp. 557–567. doi: 10.1016/j.jbusres.2020.10.030.

[2] P. Koebe and S. Bohnet-Joschko. “The Impact of Digital Transformation on Inpatient Care: Mixed Methods Study”. In: JMIR Public Health and Surveillance 9 (Apr. 2023), e40622. doi: 10.2196/40622.

[3] E. Hickmann, P. Richter, and H. Schlieter. “All together now – patient engagement, patient empowerment, and associated terms in personal healthcare”. In: BMC Health Services Research 22.1 (Sept. 2022), p. 1116. doi: 10.1186/s12913-022-08501-5.

[4] M. Panagioti, G. Richardson, N. Small, et al. “Self-management support interventions to reduce health care utilisation without compromising outcomes: a systematic review and meta-analysis”. In: BMC Health Services Research 14.1 (Dec. 2014), p. 356. doi: 10.1186/1472-6963-14-356.

[5] S. Kim, M. Park, and R. Song. “Effects of self-management programs on behavioral modification among individuals with chronic disease: A systematic review and meta-analysis of randomized trials”. In: PLOS ONE 16.7 (July 2021). Ed. by S. Spencer, e0254995. doi: 10.1371/journal.pone.0254995.

[6] Z. Chengyu, H. Xueyan, and F. Ying. “Research on disease management of chronic disease patients based on digital therapeutics: A scoping review”. In: DIGITAL HEALTH 10 (Jan. 2024), p. 20552076241297064. doi: 10.1177/20552076241297064.

[7] G. Eysenbach. “Recent advances: Consumer health informatics”. In: BMJ 320.7251 (June 2000), pp. 1713–1716. doi: 10.1136/bmj.320.7251.1713.

[8] M. Goeldner and S. Gehder. “Digital Health Applications (DiGAs) on a Fast Track: Insights From a Data-Driven Analysis of Prescribable Digital Therapeutics in Germany From 2020 to Mid-2024”. In: Journal of Medical Internet Research 26 (Aug. 2024), e59013. doi: 10.2196/59013.

[9] International Organization for Standardization. Health informatics — Personalized digital health — Digital therapeutics health software systems. Technical Report ISO/TR 11147:2023. Geneva: ISO, June 2023.

[10] C. Wang, C. Lee, and H. Shin. “Digital therapeutics from bench to bedside”. In: npj Digital Medicine 6.1 (Mar. 2023), p. 38. doi: 10.1038/s41746-023-00777-z.

[11] N. A. Patel and A. J. Butte. “Characteristics and challenges of the clinical pipeline of digital therapeutics”. In: npj Digital Medicine 3.1 (Dec. 2020), p. 159. doi: 10.1038/s41746-020-00370-8.

[12] U. Lee, G. Jung, E.-Y. Ma, et al. “Toward Data-Driven Digital Therapeutics Analytics: Literature Review and Research Directions”. In: IEEE/CAA Journal of Automatica Sinica 10.1 (Jan. 2023), pp. 42–66. doi: 10.1109/JAS.2023.123015.

[13] M. Cucciniello, F. Petracca, O. Ciani, and R. Tarricone. “Development features and study characteristics of mobile health apps in the management of chronic conditions: a systematic review of randomised trials”. In: npj Digital Medicine 4.1 (Oct. 2021), p. 144. doi: 10.1038/s41746-021-00517-1.

[14] K. J. Thomas Craig, L. C. Morgan, C.-H. Chen, et al. “Systematic review of context-aware digital behavior change interventions to improve health”. In: Translational Behavioral Medicine 11.5 (May 2021), pp. 1037–1048. doi: 10.1093/tbm/ibaa099.

[15] C. Pinder, J. Vermeulen, B. R. Cowan, and R. Beale. “Digital Behaviour Change Interventions to Break and Form Habits”. In: ACM Transactions on Computer-Human Interaction 25.3 (June 2018), pp. 1–66. doi: 10.1145/3196830.

[16] Y. Zhu, Y. Long, H. Wang, et al. “Digital Behavior Change Intervention Designs for Habit Formation: Systematic Review”. In: Journal of Medical Internet Research 26 (May 2024), e54375. doi: 10.2196/54375.

[17] S. Michie, M. Richardson, M. Johnston, et al. “The Behavior Change Technique Taxonomy (v1) of 93 Hierarchically Clustered Techniques: Building an International Consensus for the Reporting of Behavior Change Interventions”. In: Annals of Behavioral Medicine 46.1 (Aug. 2013), pp. 81–95. doi: 10.1007/s12160-013-9486-6.

[18] D. G. Schwartz, S. Spitzer, M. Khalemsky, et al. “Apps don’t work for patients who don’t use them: Towards frameworks for digital therapeutics adherence”. In: Health Policy and Technology 13.2 (June 2024), p. 100848. doi: 10.1016/j.hlpt.2024.100848.

[19] S. Pohlmann, A. Kunz, D. Ose, et al. “Digitalizing Health Services by Implementing a Personal Electronic Health Record in Germany: Qualitative Analysis of Fundamental Prerequisites From the Perspective of Selected Experts”. In: Journal of Medical Internet Research 22.1 (Jan. 2020), e15102. doi: 10.2196/15102.

[20] L. Schmidt, M. Pawlitzki, B. Y. Renard, S. G. Meuth, and L. Masanneck. “The three-year evolution of Germany’s Digital Therapeutics reimbursement program and its path forward”. In: npj Digital Medicine 7.1 (May 2024), p. 139. doi: 10.1038/s41746-024-01137-1.

[21] A. Prodan, L. Deimel, J. Ahlqvist, et al. “Success Factors for Scaling Up the Adoption of Digital Therapeutics Towards the Realization of P5 Medicine”. In: Frontiers in Medicine 9 (Apr. 2022), p. 854665. doi: 10.3389/fmed.2022.854665.

[22] J. S. Hong, C. Wasden, and D. H. Han. “Introduction of digital therapeutics”. In: Computer Methods and Programs in Biomedicine 209 (Sept. 2021), p. 106319. doi: 10.1016/j.cmpb.2021.106319.

[23] D. Fürstenau, M. Gersch, and S. Schreiter. “Digital Therapeutics (DTx)”. In: Business & Information Systems Engineering 65.3 (June 2023), pp. 349–360. doi: 10.1007/s12599-023-00804-z.

[24] K. Sippli, S. Deckert, J. Schmitt, and M. Scheibe. “Healthcare effects and evidence robustness of reimbursable digital health applications in Germany: a systematic review”. In: npj Digital Medicine 8.1 (Aug. 2025), p. 495. doi: 10.1038/s41746-025-01879-6. (Visited on 11/16/2025).

[25] B. Freitag, M. Uncovska, S. Meister, C. Prinz, and L. Fehring. “Cost-effectiveness analysis of mHealth applications for depression in Germany using a Markov cohort simulation”. In: npj Digital Medicine 7.1 (Nov. 2024), p. 321. doi: 10.1038/s41746-024-01324-0.

[26] M. Uncovska, B. Freitag, S. Meister, and L. Fehring. “Patient Acceptance of Prescribed and Fully Reimbursed mHealth Apps in Germany: An UTAUT2-based Online Survey Study”. In: Journal of Medical Systems 47.1 (Jan. 2023), p. 14. doi: 10.1007/s10916-023-01910-x.

[27] A. Carrera, S. Manetti, and E. Lettieri. “Rewiring care delivery through Digital Therapeutics (DTx): a machine learning-enhanced assessment and development (MLEAD) framework”. In: BMC Health Services Research 24.1 (Feb. 2024), p. 237. doi: 10.1186/s12913-024-10702-z.

[28] H. Yao, Z. Liao, X. Zhang, et al. “A comprehensive survey of the clinical trial Landscape on digital therapeutics”. In: Heliyon 10.16 (Aug. 2024), e36115. doi: 10.1016/j.heliyon.2024.e36115.

[29] M. Blanchard. “User experience research in the development of digital health products: Research letter”. In: Health Policy and Technology 12.2 (June 2023), p. 100753. doi: 10.1016/j.hlpt.2023.100753.

[30] Y.-C. Seo, S. Y. Yong, W. W. Choi, and S. H. Kim. “Meta-Analysis of Studies on the Effects of Digital Therapeutics”. In: Journal of Personalized Medicine 14.2 (Jan. 2024), p. 157. doi: 10.3390/jpm14020157.

[31] L. Harst, L. Otto, P. Timpel, et al. “An empirically sound telemedicine taxonomy – applying the CAFE methodology”. In: Journal of Public Health 30.11 (Nov. 2022), pp. 2729–2740. doi: 10.1007/s10389-021-01558-2.

[32] T. Schoormann, F. Möller, and D. Szopinski. “Exploring Purposes of Using Taxonomies”. In: Wirtschaftsinformatik 2022 Proceedings. Issue: 5. 2022.

[33] S. N. Weimar, H. Minaschek, R. S. Martjan, and O. Terzidis. “Decoding Digital Therapeutics: A Qualitative Analysis of Software Features in German Digital Health Applications”. In: Studies in Health Technology and Informatics. Ed. by E. Andrikopoulou, P. Gallos, T. N. Arvanitis, et al. IOS Press, May 2025. doi: 10.3233/SHTI250507.

[34] R. C. Nickerson, U. Varshney, and J. Muntermann. “A method for taxonomy development and its application in information systems”. In: European Journal of Information Systems 22.3 (May 2013), pp. 336–359. doi: 10.1057/ejis.2012.26.

[35] O. Perski, A. Blandford, R. West, and S. Michie. “Conceptualising engagement with digital behaviour change interventions: a systematic review using principles from critical interpretive synthesis”. In: Translational Behavioral Medicine 7.2 (June 2017), pp. 254–267. doi: 10.1007/s13142-016-0453-1.

[36] BfArM. DiGA Verzeichnis. 2025. url: https://diga.bfarm.de/de (visited on 10/29/2025).

[37] J. L. Mair, A. Salamanca-Sanabria, M. Augsburger, et al. “Effective Behavior Change Techniques in Digital Health Interventions for the Prevention or Management of Non-communicable Diseases: An Umbrella Review”. In: Annals of Behavioral Medicine 57.10 (Sept. 13, 2023), pp. 817–835. doi: 10.1093/abm/kaad041.

[38] M. J. Page, J. E. McKenzie, P. M. Bossuyt, et al. “The PRISMA 2020 statement: an updated guideline for reporting systematic reviews”. In: BMJ (Mar. 2021), n71. doi: 10.1136/bmj.n71.

[39] J. Webster and R. T. Watson. “Analyzing the Past to Prepare for the Future: Writing a Literature Review”. In: MIS Quarterly 26.2 (2002). Publisher: Management Information Systems Research Center, University of Minnesota, pp. xiii–xxiii.

[40] D. Kundisch, J. Muntermann, A. M. Oberländer, et al. “An Update for Taxonomy Designers: Methodological Guidance from Information Systems Research”. In: Business & Information Systems Engineering 64.4 (Aug. 2022), pp. 421–439. doi: 10.1007/s12599-021-00723-x.

[41] M. Heumann, T. Kraschewski, O. Werth, and M. H. Breitner. “Reassessing taxonomy-based data clustering: Unveiling insights and guidelines for application”. In: Decision Support Systems 187 (Dec. 2024), p. 114344. doi: 10.1016/j.dss.2024.114344.

[42] BfArM. DiGA-Leitfaden (Stand: 28.12.2023, Version 3.5). 2023. url: https://www.bfarm.de/SharedDocs/Downloads/DE/Medizinprodukte/diga_leitfaden.html (visited on 10/29/2025).

[43] F. Möller, M. Stachon, C. Azkan, T. Schoormann, and B. Otto. “Designing business model taxonomies – synthesis and guidance from information systems research”. In: Electronic Markets 32.2 (June 2022), pp. 701–726. doi: 10.1007/s12525-021-00507-x.

[44] G. Strobel, L. Banh, F. Möller, and T. Schoormann. “Exploring Generative Artificial Intelligence: A Taxonomy and Types”. In: Proceedings of the Hawaii International Conference on System Sciences 2024 (HICSS-57). Hawaii International Conference on System Sciences. 2024. doi: 10.24251/HICSS.2024.546.

[45] DigaDocs. DigaDocs: Unabhängige Informationen zu Digitalen Gesundheitsanwendungen (DiGA). 2025. url: https://digadocs.de (visited on 10/29/2025).

[46] M. L. McHugh. “Interrater reliability: the kappa statistic”. In: Biochemia medica 22.3 (2012). Publisher: Hrvatsko društvo za medicinsku biokemiju i laboratorijsku medicinu, pp. 276–282.

[47] S. Lê, J. Josse, and F. Husson. “FactoMineR : An R Package for Multivariate Analysis”. In: Journal of Statistical Software 25.1 (2008). doi: 10.18637/jss.v025.i01.

[48] M. Charrad, N. Ghazzali, V. Boiteau, and A. Niknafs. “NbClust : An R Package for Determining the Relevant Number of Clusters in a Data Set”. In: Journal of Statistical Software 61.6 (2014). doi: 10.18637/jss.v061.i06.

[49] P. Nadkarni. “Core Technologies: Data Mining and “Big Data””. In: Clinical Research Computing. Elsevier, 2016, pp. 187–204. doi: 10.1016/B978-0-12-803130-8.00010-5.

[50] D. Szopinski, T. Schoormann, and D. Kundisch. “BECAUSE YOUR TAXONOMY IS WORTH IT: TOWARDS A FRAMEWORK FOR TAXONOMY EVALUATION”. In: Proceedings of the 27th European Conference on Information Systems (ECIS). Stockholm & Uppsala, Sweden, June 2019.

[51] J. J. Pokorny, A. Norman, A. P. Zanesco, et al. “Network analysis for the visualization and analysis of qualitative data.” In: Psychological Methods 23.1 (Mar. 2018), pp. 169–183. doi: 10.1037/met0000129.

[52] G. Csardi and T. Nepusz. “The igraph software package for complex network research”. In: InterJournal Complex Systems (2006), p. 1695. url: https://igraph.org.

[53] V. D. Blondel, J.-L. Guillaume, R. Lambiotte, and E. Lefebvre. “Fast unfolding of communities in large networks”. In: Journal of Statistical Mechanics: Theory and Experiment 2008.10 (Oct. 1, 2008), P10008. doi: 10.1088/1742-5468/2008/10/P10008.

[54] S. Dodd, M. Clarke, L. Becker, et al. “A taxonomy has been developed for outcomes in medical research to help improve knowledge discovery”. In: Journal of Clinical Epidemiology 96 (Apr. 2018), pp. 84–92. doi: 10.1016/j.jclinepi.2017.12.020.

[55] R. Kotov, R. F. Krueger, D. Watson, et al. “The Hierarchical Taxonomy of Psychopathology (HiTOP): A dimensional alternative to traditional nosologies.” In: Journal of Abnormal Psychology 126.4 (May 2017), pp. 454–477. doi: 10.1037/abn0000258.

[56] T. Therneau and B. Atkinson. rpart: Recursive Partitioning and Regression Trees. 2025. url: https://CRAN.R-project.org/package=rpart.

[57] M. Uncovska, B. Freitag, S. Meister, and L. Fehring. “Rating analysis and BERTopic modeling of consumer versus regulated mHealth app reviews in Germany”. In: npj Digital Medicine 6.1 (June 21, 2023), p. 115. doi: 10.1038/s41746-023-00862-3.

[58] GKV-Spitzenverband. DiGA-Bericht des GKV-Spitzenverbandes-2024-. (Accessed on September 10, 2025). Berlin: GKV-Spitzenverband, Apr. 1, 2025. url: https://www.gkv-spitzenverband.de/media/dokumente/krankenversicherung_1/telematik/digitales/2024_DiGA-Bericht_final.pdf.

[59] World Health Organization. Classification of digital interventions, services and applications in health: a shared language to describe the uses of digital technology for health. World Health Organization, 2023. url: https://www.who.int/publications/i/item/9789240081949 (visited on 10/29/2025).

[60] P. Olla and C. Shimskey. “mHealth taxonomy: a literature survey of mobile health applications”. In: Health and Technology 4.4 (Apr. 2015), pp. 299–308. doi: 10.1007/s12553-014-0093-8.

[61] M. J. Hashim. “User interactivity in eHealth applications: A novel taxonomy”. In: 2016 12th International Conference on Innovations in Information Technology (IIT). Al-Ain, United Arab Emirates: IEEE, Nov. 2016, pp. 1–4. doi: 10.1109/INNOVATIONS.2016.7880023.

[62] M. Greve, T.-B. Lembcke, S. Diederich, A. B. Brendel, and L. M. Kolbe. “Healthy by App – Towards a Taxonomy of Mobile Health Applications”. In: PACIS 2020 Proceedings 217 (2020).

[63] M. Glöggler and E. Ammenwerth. “Improvement and Evaluation of the TOPCOP Taxonomy of Patient Portals: Taxonomy-Evaluation-Delphi (TED) Approach”. In: Journal of Medical Internet Research 23.10 (Oct. 2021), e30701. doi: 10.2196/30701.

[64] M. Almalki and A. Giannicchi. “Health Apps for Combating COVID-19: Descriptive Review and Taxonomy”. In: JMIR mHealth and uHealth 9.3 (Mar. 2021), e24322. doi: 10.2196/24322.

[65] E. Dhar, A. N. Bah, I. A. Chicchi Giglioli, et al. “A Scoping Review and a Taxonomy to Assess the Impact of Mobile Apps on Cancer Care Management”. In: Cancers 15.6 (Mar. 15, 2023), p. 1775. doi: 10.3390/cancers15061775.

[66] N. S. Mueller, O. Werth, C. M. Koenig, and M. H. Breitner. “How is Your Mood Today? - A Taxonomy-based Analysis of Apps for Depression”. In: AMCIS 2022 Proceedings. 2022.

[67] D. A. Greenwood, M. L. Litchman, D. Isaacs, et al. “A New Taxonomy for Technology-Enabled Diabetes Self-Management Interventions: Results of an Umbrella Review”. In: Journal of Diabetes Science and Technology 16.4 (July 2022), pp. 812–824. doi: 10.1177/19322968211036430.

[68] W. Wood and D. T. Neal. “Healthy through Habit: Interventions for Initiating & Maintaining Health Behavior Change”. In: Behavioral Science & Policy 2.1 (Apr. 2016), pp. 71–83. doi: 10.1177/237946151600200109.

[69] M. Schmidt-Kraepelin, S. Thiebes, M. C. Tran, and A. Sunyaev. “What’s in the game? Developing a taxonomy of gamification concepts for health apps”. In: Proceedings of the 51st Hawaii International Conference on System Sciences. 2018.

[70] R. Hervas, D. Ruiz-Carrasco, T. Mondejar, and J. Bravo. “Gamification mechanics for behavioral change: a systematic review and proposed taxonomy”. In: Proceedings of the 11th EAI International Conference on Pervasive Computing Technologies for Healthcare. Barcelona Spain: ACM, May 2017, pp. 395–404. doi: 10.1145/3154862.3154939.

[71] R. Krämer, L. Köhne-Volland, A. Schumacher, and S. Köhler. “Efficacy of a Web-Based Intervention for Depressive Disorders: Three-Arm Randomized Controlled Trial Comparing Guided and Unguided Self-Help With Waitlist Control”. In: JMIR Formative Research 6.4 (Apr. 2022), e34330. doi: 10.2196/34330.

[72] B. Meyer, J. Bierbrodt, J. Schröder, et al. “Effects of an Internet intervention (Deprexis) on severe depression symptoms: Randomized controlled trial”. In: Internet Interventions 2.1 (Mar. 2015), pp. 48–59. doi: 10.1016/j.invent.2014.12.003. (Visited on 11/16/2025).

[73] T. Berger, K. Hämmerli, N. Gubser, G. Andersson, and F. Caspar. “Internet-Based Treatment of Depression: A Randomized Controlled Trial Comparing Guided with Unguided Self-Help”. In: Cognitive Behaviour Therapy 40.4 (Dec. 2011), pp. 251–266. doi: 10.1080/16506073.2011.616531.

[74] J. M. Zill, E. Christalle, B. Meyer, M. Härter, and J. Dirmaier. “The Effectiveness of an Internet Intervention Aimed at Reducing Alcohol Consumption in Adults”. In: Deutsches Ärzteblatt international (Feb. 2019). doi: 10.3238/arztebl.2019.0127.

[75] A. Etzelmueller, E. Heber, H. Horvath, et al. “The Evaluation of the GET.ON Nationwide Web-Only Treatment Service for Depression- and Stress-Related Symptoms: Naturalistic Trial”. In: Journal of Medical Internet Research 26 (Feb. 2024), e42976. doi: 10.2196/42976.

[76] A. Baumeister, L. Schuurmans, and S. Moritz. “Anxiety unplugged: Effectiveness of an unguided, transdiagnostic, web-based intervention for anxiety disorders—A randomized controlled trial”. In: Internet Interventions 41 (Sept. 2025), p. 100867. doi: 10.1016/j.invent.2025.100867.

[77] S. Sharma and B. A. Kumar. “A systematic review of user-based usability testing practices in selfcare mHealth apps”. In: DIGITAL HEALTH 11 (May 2025), p. 20552076251374184. doi: 10.1177/20552076251374184.

[78] P. Weintraub, T. M. Dunn, and J. Yager. “Relationship of Behavioral Addictions to Eating Disorders and Substance Use Disorders”. In: Eating Disorders, Addictions and Substance Use Disorders. Ed. by T. D. Brewerton and A. Baker Dennis. Berlin, Heidelberg: Springer Berlin Heidelberg, 2014, pp. 405–428. doi: 10.1007/978-3-642-45378-6_18.

[79] L. Breiman, J. H. Friedman, R. A. Olshen, and C. J. Stone. Classification And Regression Trees. 1st ed. Routledge, Oct. 2017. doi: 10.1201/9781315139470.

[80] J. M. Lipschitz, R. Van Boxtel, J. Torous, et al. “Digital Mental Health Interventions for Depression: Scoping Review of User Engagement”. In: Journal of Medical Internet Research 24.10 (Oct. 2022), e39204. doi: 10.2196/39204.

[81] M. A. McVay, G. G. Bennett, D. Steinberg, and C. I. Voils. “Dose–response research in digital health interventions: Concepts, considerations, and challenges.” In: Health Psychology 38.12 (Dec. 2019), pp. 1168–1174. doi: 10.1037/hea0000805.

[82] T. R. Cohen Rodrigues, T. Reijnders, L. D. Breeman, et al. “Use Intention and User Expectations of Human-Supported and Self-Help eHealth Interventions: Internet-Based Randomized Controlled Trial”. In: JMIR Formative Research 8 (Feb. 2024), e38803. doi: 10.2196/38803.

[83] C. K. Eaton, E. McWilliams, D. Yablon, et al. “Cross-Cutting mHealth Behavior Change Techniques to Support Treatment Adherence and Self-Management of Complex Medical Conditions: Systematic Review”. In: JMIR mHealth and uHealth 12 (May 2024), e49024–e49024. doi: 10.2196/49024.

[84] R. A. Asbjørnsen, M. L. Smedsrød, L. Solberg Nes, et al. “Persuasive System Design Principles and Behavior Change Techniques to Stimulate Motivation and Adherence in Electronic Health Interventions to Support Weight Loss Maintenance: Scoping Review”. In: Journal of Medical Internet Research 21.6 (June 2019), e14265. doi: 10.2196/14265.

[85] F. Reinsch, T. G. Weimann, and J. Stark. “From Instant Cues to Fading Views: Piloting Just-in-Time and Fading Strategies in Digital Habit Formation”. In: AMCIS 2025 Proceedings. 2025.

[86] G. D. Giebel, C. Speckemeier, C. Abels, et al. “Problems and Barriers Related to the Use of Digital Health Applications: Scoping Review”. In: Journal of Medical Internet Research 25 (May 2023), e43808. doi: 10.2196/43808.

[87] Z. Galavi, S. Norouzi, and R. Khajouei. “Heuristics used for evaluating the usability of mobile health applications: A systematic literature review”. In: DIGITAL HEALTH 10 (Jan. 2024), p. 20552076241253539. doi: 10.1177/20552076241253539.

[88] J. Liang, Q. Fang, X. Jiao, et al. “Approved trends and product characteristics of digital therapeutics in four countries”. In: npj Digital Medicine 8.1 (May 2025), p. 308. doi: 10.1038/s41746-025-01660-9. (Visited on 11/16/2025).

[89] J. Schuffelen, L. F. Maurer, N. Lorenz, et al. “The clinical effects of digital cognitive behavioral therapy for insomnia in a heterogenous study sample: results from a randomized controlled trial”. en. In: SLEEP 46.11 (Nov. 2023), zsad184. doi: 10.1093/sleep/zsad184. (Visited on 11/26/2025).

[90] Z. Jiang, X. Huang, Z. Wang, et al. “Embodied Conversational Agents for Chronic Diseases: Scoping Review”. In: Journal of Medical Internet Research 26 (Jan. 2024), e47134. doi: 10.2196/47134.

[91] T. G. Weimann, H. Schlieter, and A. B. Brendel. “Virtual Coaches: Background, Theories, and Future Research Directions”. In: Business & Information Systems Engineering 64.4 (Aug. 2022), pp. 515–528. doi: 10.1007/s12599-022-00757-9.

[92] T. Weimann, H. Schlieter, F. Reinsch, and T. Ziemssen. “Are Embodied Conversational Agents effective Tools for collecting Patient-reported Outcome Measures? – Towards a novel Approach in Multiple Sclerosis Care”. In: ICIS 2022 Proceedings. 2022.

[93] D. Haag, D. Kumar, S. Gruber, et al. “The Last JITAI? Exploring Large Language Models for Issuing Just-in-Time Adaptive Interventions: Fostering Physical Activity in a Prospective Cardiac Rehabilitation Setting”. In: Proceedings of the 2025 CHI Conference on Human Factors in Computing Systems. Yokohama Japan: ACM, Apr. 2025, pp. 1–18. doi: 10.1145/3706598.3713307.

[94] O. Freyer, S. Jayabalan, J. N. Kather, and S. Gilbert. “Overcoming regulatory barriers to the implementation of AI agents in healthcare”. In: Nature Medicine (July 2025). doi: 10.1038/s41591-025-03841-1.

[95] R. AlSaad, A. Abd-alrazaq, S. Boughorbel, et al. “Multimodal Large Language Models in Health Care: Applications, Challenges, and Future Outlook”. In: Journal of Medical Internet Research 26 (Sept. 2024), e59505. doi: 10.2196/59505.

[96] T. M. Maddox, P. Embí, J. Gerhart, et al. “Generative AI in Medicine — Evaluating Progress and Challenges”. In: New England Journal of Medicine 392.24 (June 2025), pp. 2479–2483. doi: 10.1056/NEJMsb2503956.

[97] A. Willms, J. Rush, S. Hofer, R. E. Rhodes, and S. Liu. “Advancing physical activity research methods using realtime and adaptive technology: A scoping review of “no-code” mobile health app research tools.” In: Sport, Exercise, and Performance Psychology 14.1 (Feb. 2025). Publisher: American Psychological Association (APA), pp. 250–267. doi: 10.1037/spy0000360.

[98] T. G. Weimann. “Paving the Way for the Low-/No-Code Development of Digital Therapeutics: The DTxTAPP Framework”. In: Enterprise Design, Operations, and Computing. EDOC 2023 Workshops. Ed. by T. P. Sales, S. De Kinderen, H. A. Proper, et al. Vol. 498. Series Title: Lecture Notes in Business Information Processing. Cham: Springer Nature Switzerland, 2024, pp. 265–280. doi: 10.1007/978-3-031-54712-6_16.

